# Adolescent Intelligence and Imaging-Based Atherosclerosis in middle age: A Population Study of Swedish Men

**DOI:** 10.1101/2025.10.03.25337218

**Authors:** Hampus Eriksson, Ángel Herraiz-Adillo, Patrik Wennberg, Bledar Daka, Irene Esteban-Cornejo, Cecilia Lenander, Daniel Berglind, Ulrika Müssener, Fang Fang, Carl Johan Östgren, Karin Rådholm, Pontus Henriksson, Viktor H. Ahlqvist

## Abstract

**Importance:** Childhood or adolescent intelligence has been linked to major somatic diseases and premature mortality, but the underlying pathways remain unclear.

**Objective:** To determine whether adolescent intelligence is associated with atherosclerosis in middle age and whether this association is mediated by the American Heart Association’s Life’s Essential 8 (LE8) metric of cardiovascular health markers.

**Design:** Population-based prospective cohort study.

**Setting:** Six university hospitals in Sweden.

**Participants:** Men in the Swedish CArdioPulmonary bioImage Study who underwent mandatory military conscription in adolescence.

**Exposure:** A standardized intelligence test administered at conscription (mean age, 18.3 years; standard deviation [SD], 0.5), assessing logical reasoning, spatial ability, technical skills, and verbal comprehension. Scores were standardized to a mean of 100 (SD, 15).

**Main Outcomes and Measures:** Atherosclerosis in middle age (mean age 56.6 years; SD, 3.9), assessed using coronary computed tomography angiography (coronary stenosis: 0%, 1–49% and ≥50%) and coronary artery calcium (CAC) scores (0, 1–99, ≥100 Agatston units), and presence of unilateral or bilateral carotid plaque/s via ultrasound. LE8 components assessed in SCAPIS were analyzed as potential mediators. Associations were modeled using multinomial logistic regression, and mediation was estimated using counterfactual mediation analysis.

**Results:** A total of 8,117 men were included in the analysis of coronary stenosis, 7,958 for CAC, and 9,092 for carotid plaque. Higher adolescent intelligence was inversely associated with atherosclerosis in middle age. In analyses with extended adjustments (Model 2) for coronary stenosis, each 15-point (1 SD) increase in adolescent intelligence was associated with 17% lower odds of having ≥50% coronary stenosis (OR: 0.83; 95% CI: 0.75–0.90; P-value: <0.001). An intelligence difference between 70 and 130 corresponded to a prevalence difference of 4.1 percentage points (10.3% vs 6.2%) of coronary stenosis ≥50%. Similar associations were observed for CAC scores and carotid plaque. Mediation via LE8 accounted for 41% to 68% of the association across outcomes; the only direct effect not mediated via LE8 was observed for unilateral carotid plaque.

**Conclusions:** Higher adolescent intelligence was associated with a lower burden of atherosclerosis in middle age, with a substantial proportion of the association mediated by modifiable cardiovascular health factors.

**Key Points:** **Question:** Is there a relationship between adolescent intelligence and atherosclerosis in middle age, and could this association be mediated through cardiovascular health factors?

**Findings:** In this population-based cohort of Swedish men, higher adolescent intelligence was associated with a lower burden of atherosclerosis in middle age, measured using coronary computed tomography angiography and carotid ultrasound. About half of this association was mediated by modifiable cardiovascular health factors.

**Meaning:** These findings suggest that adolescent intelligence may shape cardiovascular health decades later, and this may be due to changes in cardiovascular health factors.

## Introduction

Higher childhood and adolescent intelligence have been linked to a lower risk of major cardiovascular (CVD), respiratory, and vascular-related neurodegenerative diseases, ^1–3^ premature mortality ^3–5^ later in life. These findings have been consistent across populations and study designs, including longitudinal cohort studies, genetic analyses, and family-based investigations.^6–9^

However, few studies have examined how childhood or adolescent intelligence influence subclinical atherosclerosis later in life.^10–13^ Most studies have examined intermediate risk factors as outcomes,^14–17^ while others have focused on diagnosed conditions or mortality.^5,18^ The former approach leaves uncertainty as to whether the findings extend to hard clinical endpoints, while the latter may be confounded by diagnostic bias (e.g., differential healthcare-seeking behavior) or non-traditional pathways to mortality (e.g., intelligence having an effect on accidental mortality but not somatic disease mortality). As a result, it remains unclear whether differences in childhood or adolescent intelligence translate into behavioral patterns—such as smoking or dietary habits—that are substantial enough to influence long-term disease risk. Ideally, this question would be addressed in a unified cohort with childhood or adolescent intelligence data, mid-life measures of behavioral health factors, and standardized assessments of subclinical disease across all participants.

To address this gap, we linked historical military conscription records with contemporary data from the Swedish CArdioPulmonary bioImage Study (SCAPIS). Using coronary computed tomography angiography (CCTA) to evaluate coronary stenosis, coronary artery calcium (CAC) scoring, and carotid ultrasound to detect plaque, we examined whether adolescent intelligence is associated with atherosclerosis in middle age, and to what extent this relationship is mediated by traditional cardiovascular health factors.

## Method

### Study design

A longitudinal study was constructed by linking male participants from SCAPIS to their historical conscription records in the Swedish Military Conscription Register (SMCR). Detailed information about SCAPIS and SMCR has been published elsewhere.^19,20^ In brief, SMCR contains data on almost all Swedish men who underwent military conscription assessments around the age of 18 years. Military conscription was compulsory during the time when participants were in adolescence. The data from SMCR linked to SCAPIS using personal identification numbers.

SCAPIS is a population-based study of 50–64-year-olds conducted at six university hospitals across Sweden (Linköping, Gothenburg, Stockholm, Umeå, Uppsala, and Malmö/Lund), comprising 30,154 participants (men = 14,646) who underwent comprehensive state-of-the-art bioimaging and cardiovascular health assessments. Participants also completed detailed questionnaires on health, lifestyle, and sociodemographic factors (**Table S1**).

### Derivation of analytical sample

The inclusion criteria for this study were having participated in SCAPIS and in conscription before the age of 20 years (**Figure S1**). For the primary complete case analysis, we excluded individuals with missing data on adolescent intelligence at conscription, or covariates (body mass index (BMI) at conscription, educational level at conscription, physical fitness at conscription, age at conscription, site of conscription, year of conscription, and site of SCAPIS). To maximize the number of participants included, we excluded participants with missing data separately for each outcome, resulting in the following participant numbers: 8,117 for the analysis of CCTA, 7,958 for the analysis of CAC, and 9,092 for the analysis of carotid plaque.

### Adolescent intelligence

The young men (<20 years) underwent a series of cognitive and physical assessments during conscription to evaluate their suitability for military service and appropriate role placement.^20,21^ The cognitive assessment comprised four standardized paper-and-pen tests evaluating: logical reasoning, spatial ability, technical skills, and verbal comprehension. The scores from these tests were summed (unweighted) to produce a total cognitive score, which was then standardized to a normal distribution within each conscription year to account for temporal differences and enable comparisons across conscription waves. To enhance comparability with other studies, we centered the distribution to a mean of 100 with a standard deviation (SD) of 15. While the exact content of the sub-tests remains confidential due to military regulations, they have been widely utilized in cognitive research and have demonstrated strong construct validity in measuring latent general intelligence.^21^ Additionally, they have been shown to correlate strongly with results on the Swedish Scholastic Aptitude Test.^22^

### Atherosclerosis in middle age

Three different types of categorical outcomes were used to evaluate the grade of atherosclerosis: coronary stenosis, CAC score, and carotid plaque.^19^ First, CCTA, a highly accurate bioimaging technique, was used to assess the grade of stenosis by considering the segment with the highest level of stenosis in any of the 11 most relevant coronary artery segments (1–3, 5–7, 9, 11–13, and 17) according to Society of Cardiovascular Computed Tomography guidelines.^23^ For CCTA, imaging was performed using a computed tomography (CT) with a Stellar Detector and dual-source (Siemens, Forcheim, Germany). Coronary stenosis was graded as follows: 0%, 1-49% or ≥50% stenosis since ≥50% stenosis in the proximal segment increased the prognostic value of mortality during 2.3 years of follow-up.^24^ Calcium blooming artifact on the CCTA was classified as 1-49% stenosis, while the presence of a coronary stent was classified as ≥50% stenosis. Second, CT was used to generate a CAC score based on the calcium levels in the coronary arteries.^25,26^ An international standard protocol was used to evaluate the CAC score.^27^ CT imaging for CAC was also performed using a Stellar Detector and dual-source (Siemens, Forcheim, Germany). The scores were categorized as follows: 0, 1-99 or ≥100 Agatston units. For CCTA stenosis and CAC, the analyses were restricted to participants having data in all of the four proximal coronary segments (1, 5, 6, and 11).

Third, ultrasound on carotid arteries was used to evaluate the carotid plaque/s. A Siemens Acuson S2000 (Siemens, Forcheim, Germany) with two-dimensional greyscale and a linear 9L4 transducer (Siemens, Forcheim, Germany) was used as equipment. The carotid plaque was defined according to the Mannheim consensus, exceeding >50% of the adjacent intima-media thickness, extending >0.5 mm into the lumen or an overall plaque thickness >1.5 mm.^28^ The categorization for the plaque was divided as follows: no plaque, unilateral plaque/s and bilateral plaques.

### Cardiovascular health behaviors and factors in middle age

To examine how cardiovascular health behaviors and factors mediate the association between adolescent intelligence and middle age atherosclerosis, we applied the American Heart Association’s Life’s Essential 8 (LE8) framework. This composite index includes eight health components including four health behaviors (physical activity, diet, sleep health, and smoking habits) and four health factors (blood pressure [BP], BMI, blood cholesterol, and blood glucose). Medication used for high BP, blood cholesterol, and blood glucose groups was also considered. The LE8 score ranges from 0 to 100 (ideal cardiovascular health). All components were assessed at the time of SCAPIS participation.^29,30^ Details of the LE8 scoring and each component are provided in **Tables S2**.^30^

### Covariates

A directed acyclic graph (DAG) was constructed to identify potential confounding (**Figure S2**). Covariates included: BMI measured at conscription using calibrated scales and stadiometers (kg/m²), and physical fitness at conscription assessed via knee extension strength (N)^31^ and cardiorespiratory fitness from a maximal cycle ergometer test (W).

Educational level (attained or enrolled) at conscription was classified as primary school, secondary school, or university-level education collected from conscription. To account for secular trends, year of conscription was grouped into eight two-year intervals spanning 1972–1987. Additional covariates included site at conscription, age at conscription, age at SCAPIS participation, and site at SCAPIS. Continuous covariates were modeled using both linear and cubic terms to account for potentially non-linear associations.

### Statistical analysis

Descriptive statistics are reported as means with SD or counts with percentages. Multinomial logistic regression was used to estimate covariate-adjusted odds ratios (ORs), prevalences (100 × marginal probability), and 95% confidence intervals (CI) for each categorical outcome. Intelligence was modeled both linearly—per 15 units (equivalent to 1 SD, consistent with prior literature)—and using restricted cubic splines with knots at the 5th, 35th, 65th, and 95th percentiles.^32^ To model the adjusted prevalences of coronary stenosis across each of any the 11 most clinically relevant coronary segments, we employed a random-effects multinomial logistic regression, allowing the outcome in each segment to correlate within the individual.

Finally, to explore potential mechanisms, we conducted counterfactual mediation analyses to decompose the total effect of adolescent intelligence on atherosclerosis in middle age into direct and indirect (mediated) components, along with the mediated proportion, via traditional cardiovascular health factors. Rather than testing each mediator individually, we used total LE8, incorporating a broad set of eight health components. A mediation analysis with LE8 factors and LE8 behaviors separately were also completed.^29^ The mediation analyses were performed using subsamples of all the participants with complete data for the relevant mediator variables at SCAPIS.

Two covariate adjustment models were specified to separate different levels of adjustments. Model 1 included age at conscription, site at conscription, year of conscription, site at SCAPIS, and age at SCAPIS. Model 2 built on Model 1 by additionally adjusting for BMI at conscription, physical fitness (cardiorespiratory fitness at conscription, muscle strength at conscription) at conscription, and educational level measured at conscription. Results from Model 1 are mostly presented in the Appendix. All analyses were conducted using Stata version 18 and 19.

### Sensitivity analyses

We performed additional adjustment self-reported smoking status at conscription (recalled at SCAPIS assessment), systolic and diastolic BP at conscription, and height at conscription (i.e., the Flynn effect^33^), among the subsample with available data. Due to the high number of participants with unknown education levels at conscription, we excluded those with missing in a sensitivity analysis.

As a sensitivity analyses, participants with calcium blooming artifact were reclassified as having ≥50% rather than 1–49% stenosis, and those with coronary artery stents were excluded instead of being categorized as ≥50% stenosis. Also, BMI at conscription was excluded from Model 2 because it is not entirely clear if BMI acts as a confounder or not (i.e., its causal effect on intelligence is debated).^34^ Other analyses were performed when participants with myocardial infarction, stroke, peripheral artery disease and coronary artery bypass graft were excluded. In addition, adjusted prevalences were calculated using only participants with complete data for all 11 coronary segments. Finally, an analysis was performed adjusting for educational level at SCAPIS.

## Results

### Descriptive characteristics

There were 8,117, 7,958 and 9,092 participants available for the analyses in coronary stenosis, CAC and carotid plaque, respectively. At conscription, the 8,117 adolescents with coronary stenosis data had a mean (SD) age of 18.3 (0.5) years, an intelligence score of 100.5 (14.7), a BMI of 21.2 (2.3) kg/m^2^, a knee strength of 556.9 N (113.2), and a cardiorespiratory fitness of 258.8 W (42.0). Educational level was distributed as primary (17.9%), secondary (64.7%), university (0.3%), and unknown (17.2%) (**Table 1; Table S3; Table S4**).

**Table 1.** Descriptive characteristics of the participants included in the analyses of coronary stenosis, coronary artery calcium, and carotid plaque

| Characteristics | Coronary stenosis sample | CAC sample | Carotid plaque sample |
| --- | --- | --- | --- |
| **At conscription (baseline)** | 8,117 (100.0%) | 7,958 (100.0%) | 9,092 (100.0%) |
| Adolescent intelligence | 100.5 (14.7) | 100.5 (14.7) | 100.3 (14.8) |
| Age (years) | 18.3 (0.5) | 18.3 (0.5) | 18.3 (0.5) |
| Height (cm) | 179.8 (6.4) | 179.8 (6.4) | 179.7 (6.5) |
| Weight (kg) | 68.7 (8.9) | 68.8 (8.9) | 68.9 (9.0) |
| BMI (kg/m<sup>2</sup>) | 21.2 (2.3) | 21.3 (2.3) | 21.3 (2.4) |
| Smoking status |  |  |  |
| Current smoker | 2,167 (26.7%) | 2,111 (26.5%) | 2,453 (27.0%) |
| Non-smoker | 5,694 (70.1%) | 5,597 (70.3%) | 6,324 (69.6%) |
| Unknown | 256 (3.2%) | 250 (3.1%) | 315 (3.5%) |
| Systolic BP (mmHg) | 127.8 (10.6) | 127.8 (10.6) | 127.9 (10.7) |
| Diastolic BP (mmHg) | 67.9 (9.5) | 67.9 (9.5) | 68.0 (9.5) |
| Cardiorespiratory fitness (W) | 258.8 (42.0) | 258.9 (42.0) | 258.3 (41.9) |
| Knee strength (N) | 556.9 (113.2) | 557.0 (113.3) | 556.6 (113.7) |
| Education level |  |  |  |
| Primary school | 1,454 (17.9%) | 1,419 (17.8%) | 1,652 (18.2%) |
| Secondary school | 5,248 (64.7%) | 5,151 (64.7%) | 5,855 (64.4%) |
| University degree | 22 (0.3%) | 22 (0.3%) | 29 (0.3%) |
| Unknown | 1,393 (17.2%) | 1,366 (17.2%) | 1,556 (17.1%) |
| **At SCAPIS (follow-up)** |  |  |  |
| Follow-up (years) | 38.3 (3.7) | 38.3 (3.7) | 38.3 (3.8) |
| Age (years) | 56.6 (3.9) | 56.6 (3.9) | 56.6 (3.9) |

Abbreviations: BMI, body mass index; BP, blood pressure; CAC, coronary artery calcium; N, newton; SCAPIS, Swedish CardioPulmonary bioImage Study; SD, standard deviations; W, watt.

At SCAPIS recruitment, mean age of participants was 56.6 (3.9) years. At that time, 3,872 (47.7%) participants presented with 0% coronary stenosis, 3,590 (44.2%) with 1-49% coronary stenosis and 655 (8.1%) with ≥50% coronary stenosis. According to CAC, 3,960 (49.8%) had 0 Agatston units, 2,747 (34.5%) had 1-99 Agatston units and 1,251 (15.7%) had ≥100 Agatston units. Finally, 3,714 (40.8%) had no plaque, 2,789 (30.7%) had unilateral plaque/s and 2,589 (28.5%) had bilateral plaques **(Table 2)**.

**Table 2.** Association of adolescent intelligence (per 15-unit [1 SD] increase) with coronary stenosis, coronary artery calcium, and carotid plaques in middle age, estimated using multinomial logistic regression

| Outcomes | Number of participants | Per 15-unit (1 SD) higher adolescent intelligence |  |  |  |  |  |
| --- | --- | --- | --- | --- | --- | --- | --- |
|  |  | Model 1 |  |  | Model 2 |  |  |
| Coronary stenosis | 8,117 (100.0%) | OR | 95% CI | P | OR | 95% CI | P |
| 0% stenosis | 3,872 (47.7%) | 1.00 | – | – | 1.00 | – | – |
| 1–49% stenosis | 3,590 (44.2%) | 0.90 | 0.86–0.94 | <0.001 | 0.91 | 0.87–0.96 | <0.001 |
| ≥50% stenosis | 655 (8.1%) | 0.79 | 0.72–0.86 | <0.001 | 0.83 | 0.75–0.90 | <0.001 |
| Coronary artery calcium | 7,958 (100.0%) |  |  |  |  |  |  |
| 0 Agatston units | 3,960 (49.8%) | 1.00 | – | – | 1.00 | – | – |
| 1–99 Agatston units | 2,747 (34.5%) | 0.91 | 0.87–0.96 | <0.001 | 0.93 | 0.88–0.98 | 0.008 |
| ≥100 Agatston units | 1,251 (15.7%) | 0.87 | 0.81–0.93 | <0.001 | 0.90 | 0.83–0.96 | 0.003 |
| Carotid plaque | 9,092 (100.0%) |  |  |  |  |  |  |
| No plaque | 3,714 (40.8%) | 1.00 | – | – | 1.00 | – | – |
| Unilateral plaque/s | 2,789 (30.7%) | 0.97 | 0.92–1.02 | 0.218 | 0.96 | 0.91–1.01 | 0.140 |
| Bilateral plaques | 2,589 (28.5%) | 0.87 | 0.82–0.91 | <0.001 | 0.88 | 0.83–0.93 | <0.001 |

Model 1 adjusts for age at conscription, site at conscription, year at conscription, site at SCAPIS, age at SCAPIS. Model 2 builds on Model 1 by additionally adjusting for BMI at conscription, educational level at conscription and physical fitness at conscription.
Abbreviations: BMI, body mass index; CI, confidence interval; OR, odds ratio; SCAPIS, Swedish CARDioPulmonary bioImage Study; SD, standard deviations.

### Adolescent intelligence and atherosclerosis

After adjusting for covariates, higher adolescent intelligence was inversely associated with measures of atherosclerosis in middle age (**Table 2**). In Model 2, each 15-point (1 SD) higher intelligence was associated with 9% lower odds of having 1-49% coronary stenosis (OR: 0.91; 95% CI: 0.87–0.96), and 17% lower odds of having ≥50% coronary stenosis (OR: 0.83; 95% CI: 0.75–0.90). Similar associations were observed for the other two outcomes: higher intelligence was linked to 10% lower odds of having a CAC score ≥100 Agatston units (OR: 0.90; 95% CI: 0.83–0.96) and 12% lower odds of bilateral carotid plaques (OR: 0.88; 95% CI: 0.83–0.93).

Findings were consistent when modeling adolescent intelligence as a non-linear exposure using restricted cubic splines (**Figure 1; Figure S3**). In the adjusted Model 2, results for both coronary stenosis and CAC demonstrated a graded, inverse association, with higher adolescent intelligence linked to progressively lower levels of atherosclerosis in middle age. The inverse association appeared more pronounced for severe outcomes, particularly for coronary stenosis ≥50% and CAC ≥100 Agatston units. Associations with carotid plaque/s appeared generally weaker than those observed for coronary measures.

**Figure 1.**
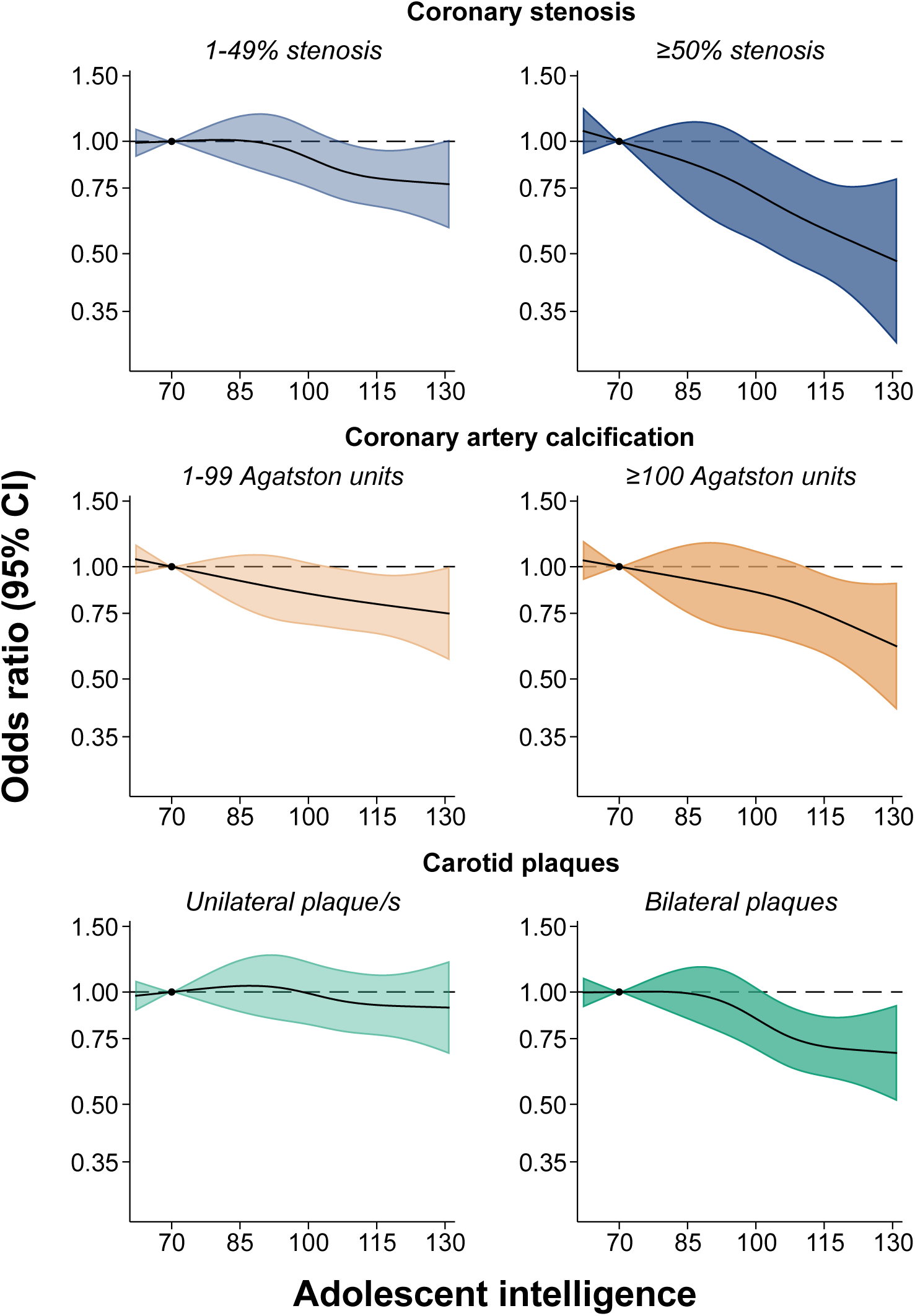
Odds ratios and 95% confidence intervals for the association between adolescent intelligence and coronary stenosis, coronary artery calcium, and carotid plaque/s in middle age, modeled with restricted cubic splines. The reference value (dot) is an adolescent intelligence of 70 (2 SD below the mean). Estimates are from Model 2, age at conscription, site at conscription, year at conscription, site at SCAPIS, age at SCAPIS, BMI at conscription, educational level at conscription and physical fitness at conscription. Abbreviations: BMI, body mass index; CI, confidence intervals; OR, odds ratio; SCAPIS, Swedish CArdioPulmonary bioImage Study.

### Absolute risk across the 11 most relevant coronary artery segments

In general, individuals with lower adolescent intelligence had a higher adjusted prevalence of coronary atherosclerosis across segments compared to those with higher intelligence (**Figure 2; Table S5; Table S6**).

**Figure 2.**
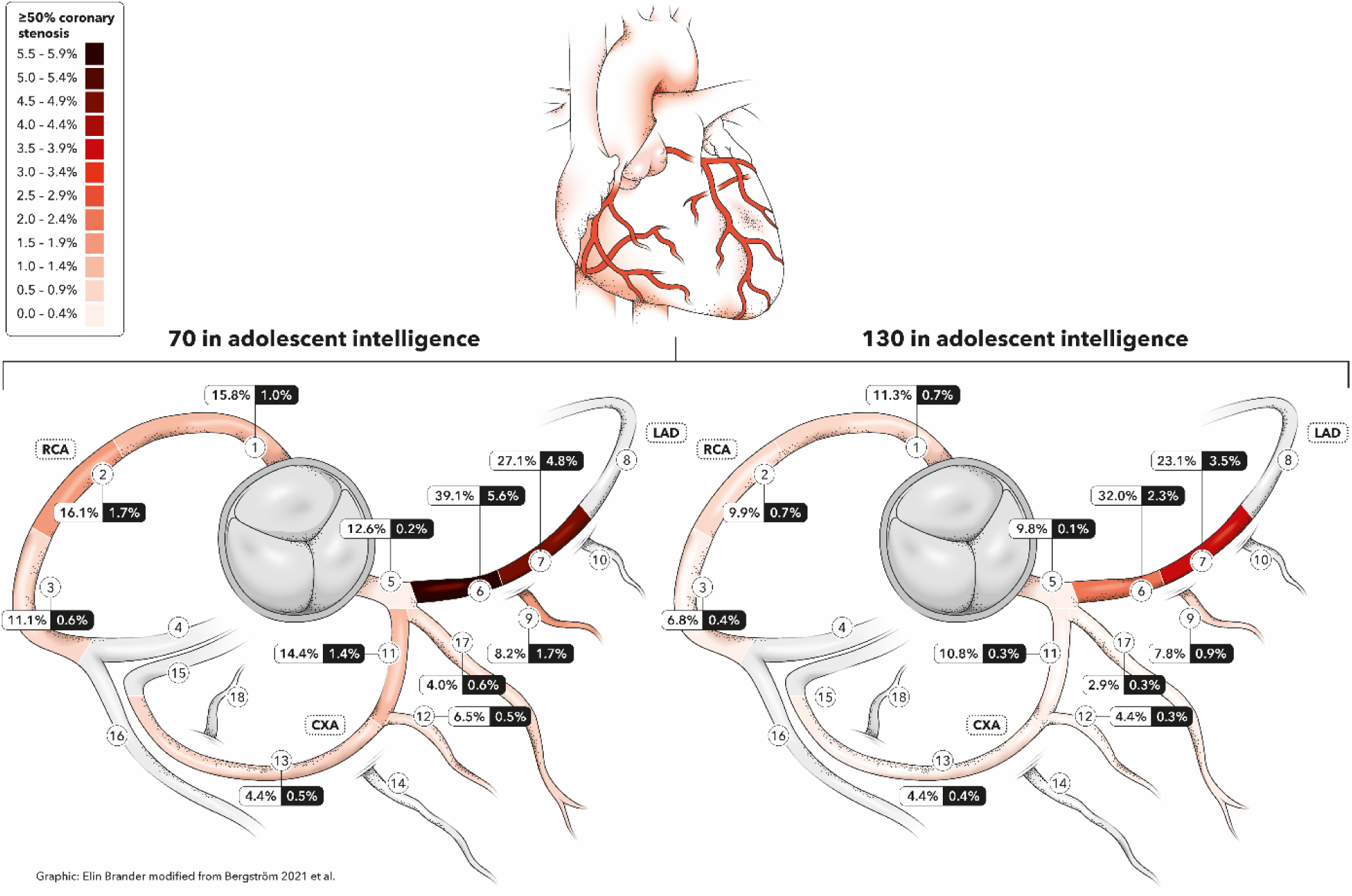
Adjusted prevalences of coronary stenosis detected by CCTA across the 11 most relevant coronary artery segments in middle age, stratified by adolescent intelligence (IQ 70 vs. 130). Estimates are from Model 2, which is adjusted for age at conscription, site at conscription, year at conscription, site at SCAPIS, age at SCAPIS, BMI at conscription, educational level at conscription and physical fitness at conscription. Coronary artery segments are defined according to the Society of Cardiovascular Computed Tomography guidelines.^40^ White and black boxes show adjusted prevalences of coronary stenosis 1–49% and ≥50%, respectively. Abbreviations: CCTA, coronary computed tomography angiography; CXA, Circumflex coronary artery; LAD, Left anterior descending coronary artery; RCA, Right coronary artery; SCAPIS, Swedish CArdioPulmonary bioImage Study.

Overall, an intelligence difference between 70 and 130 corresponded to a 5.7–percentage point difference in the prevalence of 1–49% coronary stenosis (47.0% vs 41.3%) and a 4.1–percentage point difference in the prevalence of ≥50% stenosis (10.3% vs 6.2%). Similar associations were observed for CAC scores and carotid plaque.

### Mediation through traditional cardiovascular health factors

The percentage of the association mediated through the total LE8 score measured at SCAPIS was 48% for severe coronary stenosis (≥50%), 68% for CAC (≥100 Agatston units), and 41% for bilateral carotid plaques (**Figure 3; Table S7**). The mediated proportion was somewhat larger for LE8 factors rather than for LE8 behaviors (**Table S7**).

**Figure 3.**
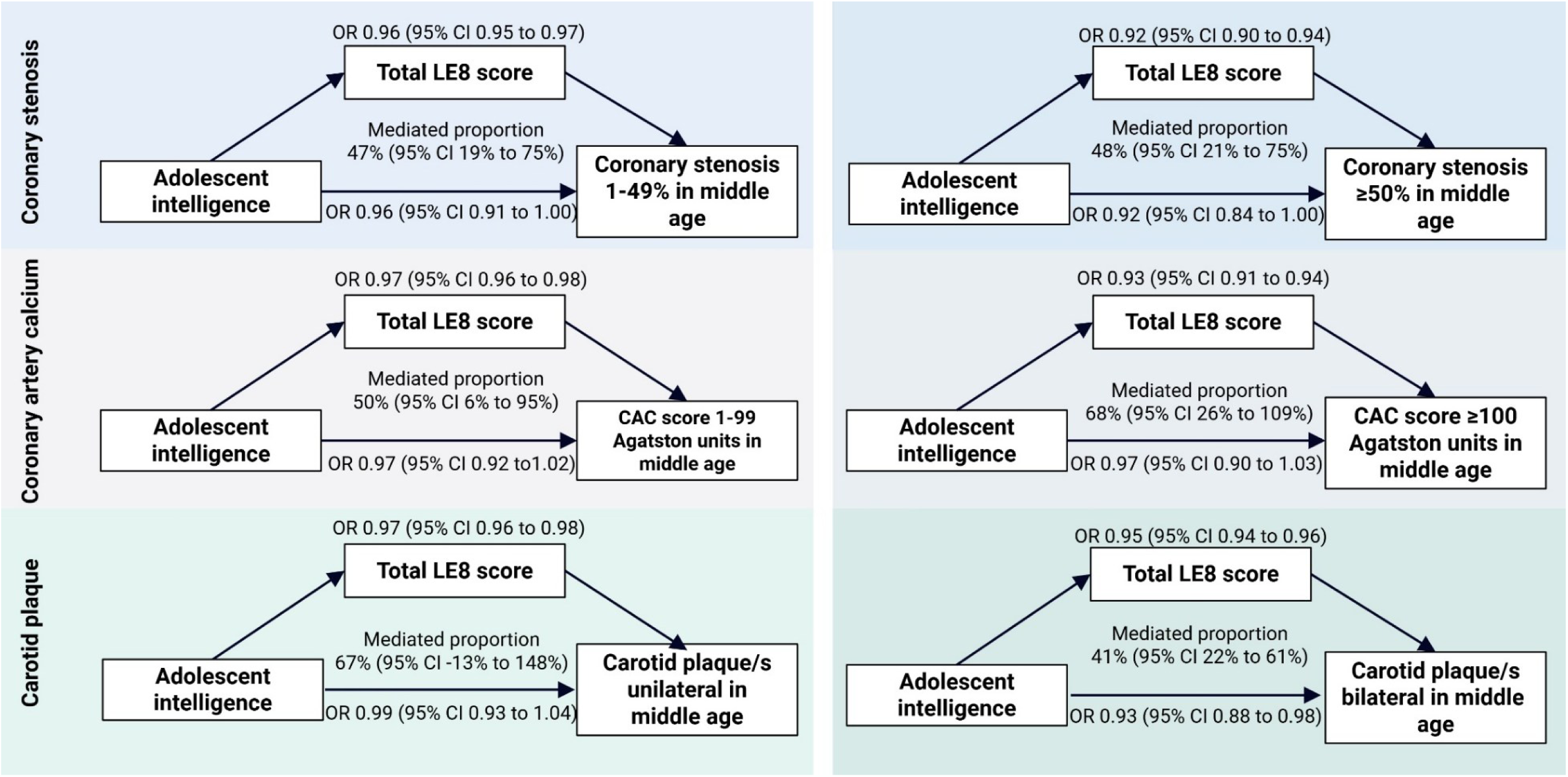
Direct and indirect effects of adolescent intelligence on coronary stenosis, coronary artery calcium, and carotid plaques in middle age, mediated via Life’s Essential 8 from SCAPIS, estimated using counterfactual mediation analysis. The LE8 variable included physical activity, diet, sleep health, and smoking habits, blood pressure, BMI, blood cholesterol, and blood glucose from SCAPIS. Mediated proportions are presented as percentages (note this estimate and its 95% CI is not bounded between 0-100). Estimates are from Model 2, which is adjusted for age at conscription, site at conscription, year at conscription, site at SCAPIS, age at SCAPIS, BMI at conscription, educational level at conscription and physical fitness at conscription. Sample sizes were 7,905 participants for coronary stenosis, 7,915 participants for CAC and 8,117 participants for carotid plaque. Abbreviations: BMI, Body Mass Index; CAC, coronary artery calcium; CI, confidence interval; LE8, Life’s Essential 8; OR, odds ratio; SCAPIS, Swedish CArdioPulmonary bioImage Study.

### Sensitivity analyses

Results were unchanged when excluding participants with missing education, adjusting for smoking, height, and blood pressure, reclassifying blooming artifacts and excluding stents, omitting BMI, excluding prior cardiovascular disease, or adjusting for education at SCAPIS (**Tables S8–S13**). Restricting analyses to participants with complete data across all 11 coronary segments yielded similar estimates (**Table S14**).

## Discussion

In this population-based cohort study of Swedish men, with nearly four decades of follow-up, higher adolescent intelligence was associated with a lower burden of atherosclerosis in middle age, demonstrating a clear dose-response relationship. The associations were particularly pronounced for coronary stenosis and CAC. For instance, each 15-unit increase (1 SD) in adolescent intelligence was linked to 17% lower odds of coronary stenosis (≥50%). The adjusted prevalences for coronary stenosis (≥50%) were 10.3% in participants with 70 in adolescent intelligence compared with 6.2% in participants with 130 in adolescent intelligence in atherosclerosis. Finally, a substantial proportion of the association between adolescent intelligence and middle age atherosclerosis was mediated by cardiovascular health (measure as total LE8 score), particularly cardiovascular health factors.

### Comparisons with existing literature

To our knowledge, only four prior studies have investigated the link between childhood or adolescent intelligence and subclinical markers of CVD later in life.^10–13^ In another Swedish study of 1,009 men, lower scores on conscription adolescent intelligence tests were associated with increased risk of carotid plaques, an association that appeared to be mediated by socioeconomic status and lifestyle habits.^11^ A U.S study of 4,286 male veterans found that lower intelligence in late adolescence was associated with a lower ankle–brachial index (ABI) at 40.^35^ Another study, based on a smaller UK cohort (n = 412), reported that higher intelligence at age 11 was associated with lower carotid intima–media thickness in adulthood.^12^ However, a separate UK study with longer followup found no differences when studying the same childhood intelligence measures and carotid intima-media thickness, carotid stenosis >25%, or ABI <0.9 at age 73.^13^ While informative, these prior studies were limited by relatively small sample sizes and reliance on indirect or surrogate markers of vascular health. In contrast, CCTA provides high-resolution, direct visualization of coronary stenosis and plaque phenotype—including calcified, non-calcified, and mixed plaques. Our findings therefore extend these prior works by offering a more comprehensive, imaging-based assessment of atherosclerosis. In addition, this study demonstrates that cardiovascular health factors mediate the association between adolescent intelligence and atherosclerosis in middle age.

### Strength and limitations

Our study has several notable strengths. First, we used high-resolution CCTA, which allows for precise quantification of coronary stenosis and the characterization of both calcified and non-calcified plaques. Second, the nearly 40-year follow-up provides a rare opportunity to examine long-term associations from adolescence into late middle age. Third, the study includes a large, well-characterized cohort, addressing the limitations of small sample sizes that have affected prior research in this area.^10,12,13,36^

Several limitations should also be considered. First, the study sample included only men, limiting generalizability to women. Albeit we might speculate that any potential mechanism may be similar in women, it should be noted that men tend to develop cardiovascular events and atherosclerosis—partly due to less favorable cardiovascular risk profiles.^37,38^ Second, as with any observational study, there may always be the risk of residual confounding. Third, although long follow-up is a strength, the lack of repeated imaging on atherosclerosis over time limits our ability to pinpoint *when* differences in atherosclerosis emerge. Fourth, the mediators were assessed contemporaneously with the outcome. This raises the possibility of reverse causation for the mediators, where individuals with atherosclerosis—although asymptomatic (i.e., no clinically diagnosed CVD)—may have modified their behaviors in response to prior medical evaluations or advice. Future studies incorporating repeated assessments of cardiovascular health profiles across the life course would help clarify these pathways. While this study focused on cardiovascular health mediators, other factors may also mediate the association—either independently or in conjunction with the studied pathway—such as socioeconomic status and education level.^11^ Finally, although military conscription was virtually mandatory and included most men, individuals with significant functional disabilities were typically exempted. Therefore, the findings may not be generalizable to populations with such disabilities.

### Clinical implications

To our knowledge, this is the first study to investigate the relationship between adolescent intelligence and CCTA, an accurate bioimaging method for assessing atherosclerosis in middle age, both according to relative and absolute metrics. Our study helps bridge the gap between lower adolescent intelligence and increased risk of atherosclerosis, providing a potential mechanistic pathway linking cognitive ability in adolescence to later-life health outcomes.

If the association is presumed—or can be demonstrated—to be causal, its magnitude would be comparable to that of several well-established cardiovascular risk factors. For instance, each 15-point higher adolescent intelligence was associated with a 17% lower odds of coronary stenosis ≥50% (OR: 0.83; 95% CI: 0.75–0.90), which is roughly comparable to the 22% lower odds observed in adolescents in the top third of cardiorespiratory fitness compared with those in the bottom third (OR: 0.78; 95% CI: 0.61– 0.99).^39^

The adjusted prevalences highlight the potential impact of adolescent intelligence: for severe coronary stenosis, having an intelligence score of 70 versus 130 in adolescence was associated with a 4.1% higher marginal prevalence for severe coronary stenosis, compared with a 2.6% difference for low cardiorespiratory fitness and low strength versus high cardiorespiratory fitness and high strength in the same cohort.^39^

### Conclusions

In this large population-based study of Swedish men, higher adolescent intelligence was associated with lower burden of subclinical atherosclerosis in middle age, including coronary stenosis, CAC, and carotid plaque. A substantial proportion of this association appeared to be mediated through modifiable cardiovascular health factors. These findings suggest that adolescent intelligence may shape atherosclerosis burden health decades later, and this may be due to changes in cardiovascular health behaviors and factors.

## Data Availability

All data produced in the present work are contained in the manuscript and in the supplementary amterials.

## Statements and Declarations

### Funding

The Swedish Heart–Lung Foundation was the main funder of the Swedish CArdioPulmonary bioImage Study (SCAPIS). Additional funding was provided by the Knut and Alice Wallenberg Foundation, the Swedish Research Council and VINNOVA (Sweden‘s Innovation Agency), University of Gothenburg and Sahlgrenska University Hospital, Karolinska Institutet and Stockholm County Council, Linköping University and University Hospital, Lund University and Skåne University Hospital, Umeå University and University Hospital, Uppsala University and University Hospital. Additional funding for this study was obtained from the National Institutes of Health (R01NS131433) and the Joanna Cocozza Foundation for Children’s Medical Research. The funders had no role in the design of the study, data management, data analysis, interpretation of findings, and the decision to submit the article for publication.

### Conflicts of interest

No conflicts of interest were declared.

### Ethics statement

Swedish Ethical Review Authority granted ethical approval for this study with reference number 2021–06408 and 2022–04375, and informed consent was obtained at SCAPIS enrollment.

### Author Contributions

VHA, PH, AHA, KR and HE contributed to the conceptualization of the study. HE performed the analysis and drafted the original manuscript. All authors critically revised the analysis plan. AHA curated the data. All authors critically revised the manuscript for important intellectual content and contributed to the development of the study methodology. HE and AHA had full access to all the data in the study and takes responsibility for the integrity of the data and the accuracy of the data analysis. All authors read and approved of the final manuscript.

### Availability of data and material

The data used for the applied example are publicly unavailable according to regulations under Swedish law. Readers interested in obtaining the data may seek similar approvals and inquire through https://www.scapis.org/.

## Notes

### Competing Interest Statement

The authors have declared no competing interest.

### Author Declarations

Swedish Ethical Review Authority granted ethical approval for this study with reference number 2021-06408 and 2022-04375, and informed consent was obtained at SCAPIS enrollment.

