## Supplementary materials for "Adolescent Intelligence and Imaging-Based Atherosclerosis in middle age: A Population Study of Swedish Men"

### Supplementary materials with tables

**Table S1.** Extended descriptions of each parameter used in the study.

**Tables S2**. Extended description of each Life's Essential 8 components.

**Table S3**. Descriptive statistics of participants excluded due to missing data.

**Table S4**. Descriptive characteristics of the participants stratified by levels of adolescent intelligence.

**Table S5.** Multinominal logistic regression with adjusted prevalences examining the association between adolescent intelligence and coronary stenosis in middle age, considering the 11 most relevant coronary artery segments and in all 18 segments.

**Table S6** Multinominal logistic regression with adjusted prevalences examining the association between adolescent intelligence and coronary stenosis in middle age, considering the seven excluded coronary artery segments.

**Table S7.** Mediation analysis using multinomial logistic regression to examine the direct and indirect effects of adolescent intelligence on coronary stenosis, Coronary Artery Calcium and carotid plaque in middle age, mediated through total LE8 score, LE8 behaviors and LE8 factors from SCAPIS.

**Table S8.** Odds ratios for atherosclerosis in middle age associated with each 15-unit (1 SD) increase in adolescent intelligence, excluding participants with missing educational level data at conscription.

**Table S9.** Odds ratios for atherosclerosis in middle age associated with each 15-unit (1 SD) increase in adolescent intelligence, with extended adjustments for smoking, height, and blood pressure at conscription.

**Table S10.** Odds ratios for atherosclerosis in middle age associated with each 15-unit (1 SD) increase in adolescent intelligence, with extended adjustments.

**Table S11.** Odds ratios for atherosclerosis in middle age associated with each 15-unit (1 SD) increase in adolescent intelligence, according to Model 2 minus BMI at conscription.

**Table S12.** Odds ratios for atherosclerosis in middle age associated with each 15-unit (1 SD) increase in adolescent intelligence, excluding participants with myocardial infarction, stroke, peripheral artery disease or coronary artery bypass graft.

**Table S13.** Odds ratios for atherosclerosis in middle age associated with each 15-unit (1 SD) increase in adolescent intelligence, with extended adjustments for educational level at SCAPIS.

**Table S14**. Odds ratios for atherosclerosis in middle age associated with each 15-unit (1 SD) increase in adolescent intelligence, considering data in all 11 coronary artery segments.

### Supplementary materials with figures

**Figure S1.** A flowchart for the study.

**Figure S2.** Directed Acyclic Graph illustrating the association between adolescent intelligence and atherosclerosis in middle age.

**Figure S3.** Association between adolescent intelligence and coronary stenosis, CAC and carotid plaque in middle age, modeled using restricted cubic splines with multinomial logistic regression.

**Table S1. Extended descriptions of each parameter used in the study.**

| | Description |
|---|---|
| **Conscription parameters**(1) | |
| Adolescent intelligence | Four subtests measuring logical reasoning, spatial ability, technical skills, and verbal comprehension. Summarized into a total score and subsequently standardized per conscription year and normalized to mean 100 SD 15. |
| Height | Measured in centimeters using a stadiometer. |
| Weight | Measured in kilograms using a scale. |
| Smoking (recalled) | Self-reported recall at SCAPIS; derived from reported age of start of smoking. |
| Systolic & diastolic BP | Blood pressure was measured using auscultation by trained staff after 5–10 minutes of supine rest, with the cuff positioned at heart level. A single reading was taken if systolic pressure was ≤145 mmHg and diastolic between 50–85 mmHg; otherwise, a second reading was obtained and used. Values were typically rounded to the nearest even number, though rounding to 5 or 10 mmHg occasionally occurred. |
| Cardiorespiratory fitness | Measured using a maximal bicycle ergometer test following a normal electrocardiography. After cycling for 5 minutes at 60–70 rates per minute with body weight–adjusted resistance, the load increased by 25 watts each minute until exhaustion. The final measure is the maximum number of watts of resistance achieved. |
| Knee strength | Knee extension strength was assessed using a isometric dynamometer in a seated position with the knee at 90° flexion, and the maximum force in newtons was recorded. |
| Educational level | Self-reported during conscription assessment. Partially missing due to lost historical records. |
| **SCAPIS parameters**(2) | |
| Height | Stadiometer measured in centimeters. |
| Weight | Scale measured in kilograms. |
| Educational level | Self-reported during SCAPIS assessment and categorized as: unfinished primary school, primary school, secondary school, and university degree. |
| Systolic & diastolic BP | Blood pressure was measured automatically twice in each arm. The mean value was used. |

| | |
|---|---|
| Fasting blood glucose | Measured using fasting blood samples analyzed at local university hospital labs. |
| Physical activity | Measured using tri-axial accelerometers worn on the right hip during waking hours for seven consecutive days.(3) |
| Diet | Dietary habits were assessed using the web-based MiniMeal-Q questionnaire,(4,5) and subsequently scored based on an adapted version of the Mediterranean Eating Pattern for Americans containing 16 components.(6) |
| Smoking | Self-reported during health assessment. |
| Sleeping habits | Self-reported during health assessment. |
| Total cholesterol | Measured using fasting blood samples analyzed at local university hospital labs. |
| Coronary stenosis | CCTA was performed using a 100–120 kV dual-source CT scanner (Somatom Definition Flash, Siemens) with protocols tailored to heart rate, rhythm variability, calcification, and body weight. Contrast-enhanced images were reconstructed using the I26f medium-smooth algorithm and assessed for atherosclerosis with syngo.via software. Coronary atherosclerosis was classified according to Society of Cardiovascular Computed Tomography guidelines, mapping the coronary tree into 11 segments.(7) |
| Coronary Artery Calcium | CAC scoring was conducted using non-contrast, ECG-gated CT scans at 120 kV (Somatom Definition Flash, Siemens). Images were reconstructed with the B35f HeartView medium CaScore algorithm and analyzed via syngo.via calcium scoring software (Volume Wizard, Siemens) following the Agatston method.(8) |
| Carotid plaques | Carotid ultrasound was performed using a Siemens Acuson S2000 with a 9L4 transducer, following a standardized protocol. Plaques in the common, internal carotid artery, and bulb were assessed bilaterally and only participants with valid readings on both sides were included. Plaques were defined according to Mannheim consensus and classified as none, unilateral, or bilateral.(9) |

Abbreviations: BMI, body mass index; BP, blood pressure; CAC, Coronary Artery Calcium; CCTA, coronary computed tomography angiography; SCAPIS, Swedish CArdioPulmonary bioImage Study; SD, standard deviations.

**Tables S2. Extended description of each Life's Essential 8 components.** *(10)*

**Four behaviors from Life's Essential 8**

**Physical activity**
Levels of physical activity were monitored using triaxial accelerometers (ActiGraph models GT3X+, wGT3X+, and wGT3X-BT). All devices included an extension filter with low-frequency (ActiGraph LCC, Pensacola, FL, USA). During all waking hours, participants should carry the accelerometers on the right hip across a 7-day period with removal only during water-based events. (3)

Physical activity intensity was defined according to the following cut-offs: light: 200–2689 counts/minute, moderate: 2690–6166 counts/minute and Vigorous with 200–2689, 2690–6166 and ≥6167 counts/minute respectively.(11) One minute of moderate activity was equivalent to one minute, while one minute of vigorous activity was weighted as two minutes. The total physical activity score was the calculated according to follow: 0 minutes = 0 points, 1–29 minutes = 20 points, 30–59 minutes = 40 points, 60–89 minutes = 60 points, 90–119 minutes = 80 points, 120–149 minutes = 90 points, ≥150 minutes = 100 points.

**Diet**
Mediterranean Eating Pattern for Americans (MEPA) served as the basis for dietary scoring.(6) Eating habits were assessed through the online questionnaire (MiniMeal-Q).(4,5)

| **Food components** | **Scoring rule** | **Points** |
|---|---|---|
| Olive oil | Regular use of olive oil | 1 |
| Butter/cream | Use of fewer than 2 of the following: cooking butter, spread butter, or rapeseed oil–butter mix | 1 |
| Cheese | ≤ 4 servings per week | 1 |
| Green leafy vegetables | ≥ 7 servings per week | 1 |
| Other vegetables | ≥ 2 servings per day | 1 |
| Berries | ≥ 2 servings per week | 1 |
| Other fruit | ≥ 2 servings per day | 1 |
| Beans | ≥ 3 servings per week | 1 |
| Nuts | ≥ 4 servings per week | 1 |
| Whole grains | ≥ 3 servings per day | 1 |
| Meat | ≤ 3 servings per week (red meat, burgers, bacon, sausage) | 1 |
| Fish | ≥ 1 serving per week | 1 |
| Chicken | ≤ 5 servings per week | 1 |
| Sweets and pastries | ≤ 4 servings per week | 1 |
| Fast food | ≤ 4 meals per week | 1 |
| Alcohol | Men: >0 to ≤2 drinks/day<br>Women: >0 to ≤1 drink/day | 1 |

An overall diet score is calculated when at least 12 food components are available. If some food components are missing, the score is adjusted using the following formula, (16 x raw diet score)/(16-number of missing food components). The total diet score is then categorized as follows, 0.0–3.9 = 0 points, 4.0–7.9 = 25 points, 8.0–11.9 = 50 points, 12.0–14.9 = 80 points and 15.0–16.0 = 100 points.

**Sleep health**

Information on sleep was collected through a self-administered questionnaire, covering average sleep duration per night (hours), presence of breathing difficulties during sleep (self- or externally reported), sleep apnea, and use of sleep apnea treatment.
The sleep habits score was categorized as follows: <4 hours = 0 points, 4–<5 hours = 20 points, 5–<6 hours or ≥10 hours = 40 points, 6–<7 hours = 70 points, 9–<10 hours = 90 points, and 7–<9 hours = 100 points. Presence of breathing difficulties or apnea resulted in a deduction of 20 points.

**Smoking habits**

Smoking habits were assessed by self-questionnaire. Information was collected on smoking status, initiation of smoking, when they quit smoking, smoking in packs per year or household exposure from cohabitants who smoke. Participants with missing information about smoking habits were categorized to the median value. They were classified to smoking habits point as follows current smoker = 0 points, former smoker quit <1 year or current nicotine delivery system user = 25 points, former smoker quit 1–<5 years = 50 points, former smoker quit ≥5 years = 75 points, never smoker = 100 points. For participants exposed to cohabitants or who had smoked, points were deducted from the nicotine exposure score depending on duration: <10 years = –10 points, 10–20 years = –15 points, >20 years = –20 points deducted from the total smoking habits score.

**Four factors from Life's Essential 8**

ATC codes from the Swedish Prescribed Drug Register were used to extract the information about medications for high blood pressure (C02, C03, C07, C08, and C09, high blood lipids (C10) and diabetic (A10).

**Blood pressure**

Brachial systolic and diastolic pressures at rest were assessed using an automated oscillometric monitor (Omron M10-IT®, Omron Healthcare Co., Kyoto, Japan) on both arms. Participants were positioned supine, with the cuff aligned to heart level, and allowed to rest for 5 minutes before the first measurement.
Subsequent recordings were taken with at least a one-minute interval to ensure full cuff deflation. If two readings from the same arm differed by more than 10 mmHg in either systolic or diastolic values, additional assessments were performed until two consecutive results fell within ±10 mmHg. No more than four trials were conducted; if the variation persisted, the last two measurements were used.

Blood pressure was measured before the use of beta-blockers (for CCTA), or alternatively on another day if needed. For the analysis, the average reading from the arm with the higher pressure was used. Systolic and diastolic blood pressure score was classified as follows: ≥160.0 or ≥100.0 = 0 points, 140.0–159.9 or 90.0–99.9 = 25 points, 130.0–139.9 or 80.0–89.9 = 50 points, 120.0–129.9/<80.0 = 75 points, and <120.0/<80.0 = 100 points. Use of antihypertensive medication resulted in a 20-point reduction.

**Body mass index**

The formula $kg/m^2$ was used to calculate body mass index (BMI). The BMI score was categorized as follows: ≥40.0 = 0 points, 35.0–39.9 = 15 points, 30.0–34.9 = 30 points, 25.0–29.9 = 70 points, and <25.0 = 100 points.

**Blood cholesterol**

Blood samples with fasting overnight were analyzed at local university hospital laboratory (Cobas Roche® and Architect Abbott®). The total cholesterol minus high density lipoprotein was used to calculate non-high-density lipoprotein (mg/dl). The blood lipid scoring system was categorized as follows: ≥220.0 = 0 points, 190.0–219.9 = 20 points, 160.0–189.9 = 40 points, 130.0–159.9 = 60 points, and <130.0 = 100 points. If blood lipids drugs were used, a subtraction of 20 points was applied.

**Blood glucose**

Blood samples with fasting overnight were analyzed at local university hospital laboratory. (Cobas Roche® and Architect Abbott®). By default, venous plasma glucose (mg/dl) and hemoglobin A1c (HbA1C %) was used as the reference. In cases where such data were not available (n = 2703 participants), capillary whole blood was measured instead. These values were converted to plasma-equivalent concentrations using the IFCC-recommended correction factor of ×1.11. Capillary blood glucose was determined with HemoCue® Glucose 201RT analyzers, which apply the glucose dehydrogenase technique. (12)

Participants were classified as having diabetes if at least one of the following criteria was met: use of antidiabetic medication, HbA1c ≥6.50%, or fasting blood glucose (FBG) ≥126.0 mg/dl. The blood glucose score were according to follows: diabetes with HbA1c≥10.0 = 0 points, diabetes with HbA1c9.0–9.9 = 10 points, diabetes with HbA1c8.0–8.9 = 20 points, diabetes with HbA1c7.0–7.9 = 30 points, diabetes with HbA1c<7.0 = 40 points, no diabetes and FBG 100.0–125.9 (or HbA1c5.70–6.49) = 60 points, no history of diabetes and FBG <100.0 (or HbA1c<5.70) = 100 points.

**Total score of Life's Essential 8**

The scoring of Life's Essential 8 (LE8) followed the recommendations from the American Heart Association (AHA). Each of the eight components was rated on a continuous scale from 0, representing the least favorable level, to 100, representing the most favorable. In this study, the LE8 score was calculated when information was available for either seven or all eight components. This produces a value between 0 and 100. When all eight components were present, the score equaled the sum of those components divided by eight, while with seven components it equaled the sum of those components divided by seven. Furthermore, separate scores were generated for LE8 behaviors and LE8 factors. These were calculated in the same manner, using the average of the three or four available components, and corrected for any missing information as described above.

**Table S3. Descriptive characteristics of participants excluded due to missing data.**

| Characteristics | Men with data from conscription and SCAPIS | Participants with available covariate data |
|---|---|---|
| **At conscription (baseline)** | n = 12,062 (100.0%) | n = 9,162 (100.0%) |
| Adolescent intelligence | 100.0 (15.0) | 100.3 (14.8) |
| Age (years) | 18.4 (0.9) | 18.3 (0.5) |
| Height (cm) | 179.6 (6.5) | 179.7 (6.5) |
| Weight (kg) | 68.7 (9.0) | 68.9 (9.0) |
| BMI (kg/m$^2$) | 21.3 (2.4) | 21.3 (2.4) |
| Smoking status | | |
| Current smoker | 3,286 (27.2%) | 2,478 (27.0%) |
| Non-smoker | 7,791 (64.6%) | 6,355 (69.4%) |
| Unknown | 985 (8.2%) | 329 (3.6%) |
| Systolic BP (mmHg) | 127.6 (10.7) | 127.9 (10.7) |
| Diastolic BP (mmHg) | 68.4 (9.5) | 67.9 (9.5) |
| Cardiorespiratory fitness (W) | 258.9 (42.8) | 258.3 (41.9) |
| Knee strength (N) | 554.8 (113.4) | 556.5 (113.7) |
| Education level | | |
| Primary school | 1,749 (14.5%) | 1,674 (18.3%) |
| Secondary school | 6,285 (52.1%) | 5,897 (64.4%) |
| University degree | 56 (0.5%) | 29 (0.3%) |
| Unknown | 3,972 (32.9%) | 1,562 (17.0%) |
| **At SCAPIS (follow-up)** | | |
| Follow-up (years) | 39.0 (4.3) | 38.3 (3.7) |
| Age (years) | 57.6 (4.4) | 56.6 (3.9) |

Abbreviations: BMI, body mass index; BP, blood pressure; CAC, Coronary Artery Calcium; SD, standard deviations; SCAPIS, Swedish CArdioPulmonary bioImage Study.

**Table S4. Descriptive characteristics of participants stratified by levels of adolescent intelligence.**

| Characteristics | <70 | 70–<85 | 85–<100 | 100–<115 | 115–130 | >130 | Total |
|---|---|---|---|---|---|---|---|
| **At conscription (baseline)** | n = 248 (2.7%) | n = 1,154 (12.6%) | n = 2,916 (31.8%) | n = 3,302 (36.0%) | n = 1,447 (15.8%) | n = 95 (1.0%) | n = 9,162 (100.0%) |
| Age (years) | 18.2 (0.5) | 18.2 (0.5) | 18.3 (0.5) | 18.3 (0.5) | 18.4 (0.5) | 18.4 (0.4) | 18.3 (0.5) |
| Height (cm) | 178.3 (6.6) | 178.8 (6.6) | 179.2 (6.4) | 180.0 (6.4) | 181.0 (6.5) | 180.9 (5.8) | 179.7 (6.5) |
| Weight (kg) | 68.3 (11.6) | 69.2 (9.8) | 68.8 (9.3) | 68.8 (8.5) | 69.3 (8.6) | 69.4 (9.3) | 68.9 (9.0) |
| BMI (kg/m$^2$) | 21.5 (3.1) | 21.6 (2.7) | 21.4 (2.4) | 21.2 (2.3) | 21.1 (2.2) | 21.2 (2.6) | 21.3 (2.4) |
| Smoking status | | | | | | | |
| Current smoker | 108 (43.5%) | 426 (36.9%) | 925 (31.7%) | 775 (23.5%) | 232 (16.0%) | 12 (12.6%) | 2,478 (27.0%) |
| Non-smoker | 123 (49.6%) | 671 (58.1%) | 1,875 (64.3%) | 2,421 (73.3%) | 1,185 (81.9%) | 80 (84.2%) | 6,355 (69.4%) |
| Unknown | 17 (6.9%) | 57 (4.9%) | 116 (4.0%) | 106 (3.2%) | 30 (2.1%) | 3 (3.2%) | 329 (3.6%) |
| Systolic BP (mmHg) | 128.2 (10.3) | 127.9 (10.5) | 127.6 (10.7) | 127.9 (10.7) | 128.1 (10.7) | 128.1 (11.1) | 127.9 (10.7) |
| Diastolic BP (mmHg) | 68.7 (9.7) | 67.8 (9.4) | 68.1 (9.4) | 68.0 (9.6) | 67.7 (9.5) | 67.5 (9.3) | 67.9 (9.5) |
| Cardiorespiratory fitness (W) | 238.2 (38.1) | 249.9 (39.0) | 255.3 (41.3) | 261.5 (42.5) | 266.8 (41.9) | 261.8 (42.2) | 258.3 (41.9) |
| Knee strength (N) | 526.0 (117.7) | 547.4 (114.9) | 554.2 (111.9) | 560.5 (114.1) | 565.6 (112.5) | 544.9 (123.9) | 556.5 (113.7) |
| Education level | | | | | | | |
| Primary school | 124 (50.0%) | 376 (32.6%) | 595 (20.4%) | 416 (12.6%) | 155 (10.7%) | 8 (8.4%) | 1,674 (18.3%) |
| Secondary school | 85 (34.3%) | 597 (51.7%) | 1,847 (63.3%) | 2,290 (69.4%) | 1,011 (69.9%) | 67 (70.5%) | 5,897 (64.4%) |
| University degree | 0 (0.0%) | 2 (0.2%) | 8 (0.3%) | 8 (0.2%) | 10 (0.7%) | 1 (1.1%) | 29 (0.3%) |
| Unknown | 39 (15.7%) | 179 (15.5%) | 466 (16.0%) | 588 (17.8%) | 271 (18.7%) | 19 (20.0%) | 1,562 (17.0%) |
| **At SCAPIS (follow-up)** | | | | | | | |
| Follow-up (years) | 38.6 (3.6) | 38.1 (3.7) | 38.5 (3.7) | 38.3 (3.8) | 38.2 (3.8) | 39.6 (3.2) | 38.3 (3.7) |
| Age (years) | 56.8 (3.8) | 56.3 (3.8) | 56.7 (3.9) | 56.7 (3.9) | 56.6 (3.9) | 57.9 (3.3) | 56.6 (3.9) |

Abbreviations: BMI, body mass index; BP, blood pressure; CAC, coronary artery calcium; N, newton; SCAPIS, Swedish CArdioPulmonary bioImage Study; SD, standard deviations; W, watt.

**Table S5. Multinominal logistic regression with adjusted prevalences examining the association between adolescent intelligence and coronary stenosis in middle age, considering any of the 11 most relevant coronary artery segments.**

| Coronary stenosis | Adolescent intelligence | Coronary artery segments n = 8,117 | | | | | | | | | | | |
|---|---|---|---|---|---|---|---|---|---|---|---|---|---|
| | | **1** | **2** | **3** | **5** | **6** | **7** | **9** | **11** | **12** | **13** | **17** | **Total, any of the 11 segments** |
| **1–49%** | **70** | 15.8 (14.0-17.8) | 16.1 (14.2-18.2) | 11.1 (9.5-12.9) | 12.6 (11.0-14.4) | 39.1 (36.5-41.8) | 27.1 (24.7-29.6) | 8.2 (6.9-9.7) | 14.4 (12.7-16.4) | 6.5 (5.4-7.9) | 4.4 (3.5-5.5) | 4.0 (3.1-5.1) | 47.0 (44.4-49.6) |
| | **85** | 14.6 (13.5-15.7) | 14.3 (13.2-15.5) | 9.8 (9.0-10.8) | 11.8 (10.9-12.9) | 37.3 (35.6-38.9) | 26.1 (24.6-27.6) | 8.1 (7.3-9.0) | 13.4 (12.4-14.6) | 5.9 (5.2-6.7) | 4.4 (3.8-5.1) | 3.7 (3.2-4.3) | 45.7 (44.1-47.3) |
| | **100** | 13.4 (12.7-14.1) | 12.7 (11.9-13.4) | 8.7 (8.1-9.3) | 11.1 (10.5-11.8) | 35.5 (34.4-36.6) | 25.1 (24.1-26.1) | 8.0 (7.4-8.6) | 12.5 (11.8-13.2) | 5.4 (4.9-5.9) | 4.4 (4.0-4.8) | 3.4 (3.0-3.8) | 44.3 (43.3-45.4) |
| | **115** | 12.3 (11.3-13.3) | 11.2 (10.3-12.2) | 7.7 (7.0-8.6) | 10.4 (9.5-11.4) | 33.7 (32.2-35.3) | 24.1 (22.7-25.5) | 7.9 (7.1-8.7) | 11.6 (10.7-12.6) | 4.9 (4.3-5.5) | 4.4 (3.8-5.1) | 3.1 (2.7-3.7) | 42.9 (41.4-44.4) |
| | **130** | 11.3 (9.9-12.9) | 9.9 (8.6-11.3) | 6.8 (5.8-8.0) | 9.8 (8.5-11.2) | 32.0 (29.6-34.5) | 23.1 (21.0-25.4) | 7.8 (6.6-9.2) | 10.8 (9.4-12.3) | 4.4 (3.6-5.4) | 4.4 (3.5-5.5) | 2.9 (2.2-3.7) | 41.3 (38.9-43.8) |
| **≥50%** | **70** | 1.0 (0.6-1.7) | 1.7 (1.1-2.5) | 0.6 (0.3-1.0) | 0.2 (0.1-0.6) | 5.6 (4.2-7.5) | 4.8 (3.6-6.2) | 1.7 (1.1-2.6) | 1.4 (0.9-2.4) | 0.5 (0.3-0.9) | 0.5 (0.3-0.9) | 0.6 (0.3-1.1) | 10.3 (8.6-11.9) |
| | **85** | 0.9 (0.7-1.3) | 1.3 (1.0-1.7) | 0.5 (0.4-0.8) | 0.2 (0.1-0.4) | 4.5 (3.8-5.4) | 4.4 (3.7-5.3) | 1.5 (1.1-1.9) | 1.0 (0.7-1.3) | 0.5 (0.3-0.7) | 0.5 (0.3-0.7) | 0.5 (0.3-0.7) | 9.1 (8.2-10.0) |
| | **100** | 0.9 (0.7-1.1) | 1.0 (0.8-1.3) | 0.5 (0.4-0.7) | 0.2 (0.1-0.3) | 3.6 (3.1-4.2) | 4.1 (3.6-4.7) | 1.2 (1.0-1.5) | 0.6 (0.5-0.8) | 0.4 (0.3-0.6) | 0.4 (0.3-0.6) | 0.4 (0.3-0.5) | 8.0 (7.4-8.6) |
| | **115** | 0.8 (0.6-1.1) | 0.8 (0.6-1.1) | 0.5 (0.3-0.7) | 0.2 (0.1-0.3) | 2.9 (2.3-3.6) | 3.8 (3.2-4.5) | 1.1 (0.8-1.4) | 0.4 (0.3-0.7) | 0.4 (0.2-0.6) | 0.4 (0.3-0.6) | 0.3 (0.2-0.5) | 7.0 (6.2-7.8) |
| | **130** | 0.7 (0.4-1.2) | 0.7 (0.4-1.1) | 0.4 (0.2-0.8) | 0.1 (0.0-0.4) | 2.3 (1.6-3.2) | 3.5 (2.6-4.7) | 0.9 (0.6-1.4) | 0.3 (0.1-0.6) | 0.3 (0.2-0.6) | 0.4 (0.2-0.7) | 0.3 (0.1-0.6) | 6.2 (5.0-7.3) |

Adjusted prevalences are presented as percentages with 95% confidence intervals in any of the 11 most relevant coronary segments. The right column presents the total adjusted prevalences from all segments, considering participants with data from any of the 11 most relevant segments. The analyses are adjusted according to Model 2, age at conscription, site at conscription, year of conscription, site at SCAPIS, age at SCAPIS, BMI at conscription, educational level at conscription and physical fitness at conscription. The cardiac segments are defined according to Society of Cardiovascular Computed Tomography guidelines.(7)
Abbreviations: BMI, body mass index; SCAPIS, Swedish CArdioPulmonary bioImage Study.

**Table S6. Multinominal logistic regression with adjusted prevalences examining the association between adolescent intelligence and coronary stenosis in middle age, considering the seven excluded coronary artery segments.**

| Coronary stenosis | Adolescent intelligence | Coronary artery segments n = 8,117 | | | | | | |
|---|---|---|---|---|---|---|---|---|
| | | **4** | **8** | **10** | **14** | **15** | **16** | **18** |
| **1–49%** | **70** | 15.8 (14.0-17.8) | 16.1 (14.2-18.2) | 11.1 (9.5-12.9) | 12.6 (11.0-14.4) | 39.1 (36.5-41.8) | 27.1 (24.7-29.6) | 8.2 (6.9-9.7) |
| | **85** | 14.6 (13.5-15.7) | 14.3 (13.2-15.5) | 9.8 (9.0-10.8) | 11.8 (10.9-12.9) | 37.3 (35.6-38.9) | 26.1 (24.6-27.6) | 8.1 (7.3-9.0) |
| | **100** | 13.4 (12.7-14.1) | 12.7 (11.9-13.4) | 8.7 (8.1-9.3) | 11.1 (10.5-11.8) | 35.5 (34.4-36.6) | 25.1 (24.1-26.1) | 8.0 (7.4-8.6) |
| | **115** | 12.3 (11.3-13.3) | 11.2 (10.3-12.2) | 7.7 (7.0-8.6) | 10.4 (9.5-11.4) | 33.7 (32.2-35.3) | 24.1 (22.7-25.5) | 7.9 (7.1-8.7) |
| | **130** | 11.3 (9.9-12.9) | 9.9 (8.6-11.3) | 6.8 (5.8-8.0) | 9.8 (8.5-11.2) | 32.0 (29.6-34.5) | 23.1 (21.0-25.4) | 7.8 (6.6-9.2) |
| **≥50%** | **70** | 1.0 (0.6-1.7) | 1.7 (1.1-2.5) | 0.6 (0.3-1.0) | 0.2 (0.1-0.6) | 5.6 (4.2-7.5) | 4.8 (3.6-6.2) | 1.7 (1.1-2.6) |
| | **85** | 0.9 (0.7-1.3) | 1.3 (1.0-1.7) | 0.5 (0.4-0.8) | 0.2 (0.1-0.4) | 4.5 (3.8-5.4) | 4.4 (3.7-5.3) | 1.5 (1.1-1.9) |
| | **100** | 0.9 (0.7-1.1) | 1.0 (0.8-1.3) | 0.5 (0.4-0.7) | 0.2 (0.1-0.3) | 3.6 (3.1-4.2) | 4.1 (3.6-4.7) | 1.2 (1.0-1.5) |
| | **115** | 0.8 (0.6-1.1) | 0.8 (0.6-1.1) | 0.5 (0.3-0.7) | 0.2 (0.1-0.3) | 2.9 (2.3-3.6) | 3.8 (3.2-4.5) | 1.1 (0.8-1.4) |
| | **130** | 0.7 (0.4-1.2) | 0.7 (0.4-1.1) | 0.4 (0.2-0.8) | 0.1 (0.0-0.4) | 2.3 (1.6-3.2) | 3.5 (2.6-4.7) | 0.9 (0.6-1.4) |

Adjusted prevalences are presented as percentages with 95% confidence intervals. The analyses are adjusted according to Model 2, age at conscription, site at conscription, year of conscription, site at SCAPIS, age at SCAPIS, BMI at conscription, educational level at conscription and physical fitness at conscription. The cardiac segments are defined according to Society of Cardiovascular Computed Tomography guidelines.(7)

Abbreviations: BMI, body mass index; SCAPIS, Swedish CArdioPulmonary bioImage Study.

**Table S7. Mediation analysis using multinomial logistic regression to examine the direct and indirect effects of adolescent intelligence on coronary stenosis, Coronary Artery Calcium and carotid plaque in middle age, mediated through total LE8 score, LE8 behaviors and LE8 factors from SCAPIS.**

| | Total | | | Behaviors | | | Factors | | |
|---|---|---|---|---|---|---|---|---|---|
| | **OR** | **95% CI** | **P** | **OR** | **95% CI** | **P** | **OR** | **95% CI** | **P** |
| **Coronary stenosis, 1–49%** | | n = 7,905 | | | n = 7,915 | | | n = 8,117 | |
| TE | 0.92 | 0.87–0.96 | <0.001 | 0.92 | 0.88–0.97 | <0.001 | 0.91 | 0.87–0.96 | <0.001 |
| NDE | 0.96 | 0.91-1.00 | 0.070 | 0.93 | 0.88–0.97 | <0.001 | 0.94 | 0.90–0.99 | 0.020 |
| NIE | 0.96 | 0.95-0.97 | <0.001 | 0.99 | 0.99-1.00 | 0.130 | 0.97 | 0.96–0.98 | <0.001 |
| Med. Proportion | 0.47 | 0.19-0.75 | <0.001 | 0.08 | -0.03-0.18 | 0.150 | 0.35 | 0.14-0.55 | <0.001 |
| **Coronary stenosis, ≥50%** | | n = 7,905 | | | n = 7,915 | | | n = 8,117 | |
| TE | 0.84 | 0.77–0.92 | <0.001 | 0.85 | 0.77-0.93 | <0.001 | 0.84 | 0.77–0.91 | <0.001 |
| NDE | 0.92 | 0.84-1.00 | 0.050 | 0.86 | 0.79-0.94 | <0.001 | 0.90 | 0.83–0.97 | 0.010 |
| NIE | 0.92 | 0.90–0.94 | <0.001 | 0.98 | 0.97-1.00 | 0.020 | 0.93 | 0.91–0.95 | <0.001 |
| Med. proportion | 0.48 | 0.21-0.75 | <0.001 | 0.10 | 0.00-0.21 | 0.050 | 0.37 | 0.17-0.57 | <0.001 |
| **CAC, 1–99 Agatston units** | | n = 7,752 | | | n = 7,762 | | | n = 7,958 | |
| TE | 0.94 | 0.89-0.99 | 0.020 | 0.94 | 0.89–0.99 | 0.030 | 0.93 | 0.88-0.98 | 0.010 |
| NDE | 0.97 | 0.92-1.02 | 0.250 | 0.95 | 0.90-1.00 | 0.040 | 0.95 | 0.91-1.01 | 0.080 |
| NIE | 0.97 | 0.96-0.98 | <0.001 | 1.00 | 0.99-1.01 | 0.470 | 0.97 | 0.97-0.98 | <0.001 |
| Med. proportion | 0.50 | 0.06-0.95 | 0.030 | 0.05 | -0.10-0.21 | 0.480 | 0.35 | 0.08-0.63 | 0.010 |
| **CAC, ≥100 Agatston units** | | n = 7,752 | | | n = 7,762 | | | n = 7,958 | |
| TE | 0.89 | 0.84-0.96 | <0.001 | 0.90 | 0.84–0.96 | <0.001 | 0.89 | 0.83–0.95 | <0.001 |
| NDE | 0.97 | 0.90-1.03 | 0.290 | 0.92 | 0.86–0.98 | 0.020 | 0.94 | 0.89-1.01 | 0.070 |
| NIE | 0.93 | 0.91-0.94 | <0.001 | 0.98 | 0.97–0.99 | <0.001 | 0.94 | 0.93–0.96 | <0.001 |
| Med. proportion | 0.68 | 0.26-1.09 | <0.001 | 0.32 | 0.09–0.55 | 0.200 | 0.50 | 0.20-0.79 | <0.001 |
| **Carotid plaque, unilateral** | | n = 8,820 | | | n = 8,834 | | | n = 9,066 | |
| TE | 0.96 | 0.9-1.01 | 0.100 | 0.96 | 0.91-1.01 | 0.110 | 0.96 | 0.91-1.01 | 0.110 |
| NDE | 0.99 | 0.93-1.04 | 0.590 | 0.97 | 0.92-1.02 | 0.230 | 0.98 | 0.93-1.03 | 0.430 |
| NIE | 0.97 | 0.96-0.98 | 0.000 | 0.99 | 0.98-1.00 | 0.020 | 0.98 | 0.97-0.99 | 0.000 |
| Med. proportion | 0.67 | -0.13-1.48 | 0.100 | 0.25 | -0.10-0.61 | 0.160 | 0.50 | -0.13-1.13 | 0.120 |
| **Carotid plaque, bilateral** | | n = 8,820 | | | n = 8,834 | | | n = 9,066 | |
| TE | 0.89 | 0.84-0.93 | <0.001 | 0.88 | 0.84–0.93 | <0.001 | 0.89 | 0.84–0.94 | <0.001 |
| NDE | 0.93 | 0.88-0.98 | 0.010 | 0.90 | 0.85–0.95 | <0.001 | 0.92 | 0.88–0.97 | <0.001 |
| NIE | 0.95 | 0.94-0.96 | <0.001 | 0.99 | 0.98–0.99 | <0.001 | 0.96 | 0.95–0.97 | <0.001 |
| Med. proportion | 0.41 | 0.22-0.61 | <0.001 | 0.12 | 0.04-0.20 | <0.001 | 0.32 | 0.16-0.48 | <0.001 |

The results are presented as percentages (95% CI). The models are adjusted according to model 2, age at conscription, site at conscription, year of conscription, site at SCAPIS, age at SCAPIS, BMI at conscription, educational level at conscription and physical fitness at conscription. Total LE8 score included diet, physical activity, sleep health, smoking, blood pressure, BMI, cholesterol, and glucose. LE8 behaviors included diet, physical activity, sleep health, and smoking. LE8 factors included blood pressure, BMI, cholesterol, and glucose.

Abbreviations: BMI, body mass index; CAC, Coronary Artery Calcium; CI, confidence interval; LE8, Life´s Essential 8; Med., mediated; NDE, natural direct effect; NIE, natural indirect effect; OR, odds ratio; SCAPIS, Swedish CArdioPulmonary bioImage Study; TE, total effect.

**Table S8. Odds ratios for atherosclerosis in middle age associated with each 15-unit (1 SD) increase in adolescent intelligence, excluding participants with missing educational level data at conscription.**

| | Main analysis [a] | | | Sensitivity analysis excluding participants with missing educational level data at conscription [b] | | |
|---|---|---|---|---|---|---|
| | Per 15–unit (1 SD) higher adolescent intelligence | | | | | |
| **Outcomes** | **OR** | **95% CI** | **P** | **OR** | **95% CI** | **P** |
| **Coronary stenosis** | n = 8,117 | | | n = 6,724 | | |
| *0% stenosis* | 1.00 | – | – | 1.00 | – | – |
| *1–49% stenosis* | 0.91 | 0.87–0.96 | <0.001 | 0.90 | 0.85–0.96 | <0.001 |
| *≥50% stenosis* | 0.83 | 0.75–0.90 | <0.001 | 0.84 | 0.76–0.93 | 0.001 |
| **Coronary artery calcium** | n = 7,958 | | | n = 6,592 | | |
| *0 Agatston units* | 1.00 | – | – | 1.00 | – | – |
| *1–99 Agatston units* | 0.93 | 0.88–0.98 | 0.008 | 0.92 | 0.87–0.98 | 0.008 |
| *≥100 Agatston units* | 0.90 | 0.83–0.96 | 0.003 | 0.90 | 0.83–0.97 | 0.010 |
| **Carotid plaque** | n = 9,092 | | | n = 7,536 | | |
| *No plaque* | 1.00 | – | – | 1.00 | – | – |
| *Unilateral plaque/s* | 0.96 | 0.91–1.01 | 0.140 | 0.95 | 0.90–1.01 | 0.089 |
| *Bilateral plaques* | 0.88 | 0.83–0.93 | <0.001 | 0.89 | 0.83–0.94 | <0.001 |

Multinomial logistic regression analysis.
[a] Main analysis refers to Model 2, adjusted for age at conscription, site at conscription, year at conscription, site at SCAPIS, age at SCAPIS, BMI at conscription, educational level at conscription, and physical fitness at conscription.
[b] The sensitivity analysis is according to Model 2, except participants with missing educational level data at conscription.
Abbreviations: BMI, body mass index; CI, confidence interval; OR, odds ratio; SCAPIS, Swedish CArdioPulmonary bioImage Study; SD, standard deviations

**Table S9. Odds ratios for atherosclerosis in middle age associated with each 15-unit (1 SD) increase in adolescent intelligence, with extended adjustments for smoking, height, and blood pressure at conscription.**

| | Main analysis [a] | | | Sensitivity analysis with extended adjustments for smoking, height, and blood pressure at conscription [b] | | |
|---|---|---|---|---|---|---|
| | Per 15–unit (1 SD) higher adolescent intelligence | | | | | |
| **Outcomes** | **OR** | **95% CI** | **P** | **OR** | **95% CI** | **P** |
| **Coronary stenosis** | | n = 8,117 | | | n = 7,853 | |
| *0% stenosis* | 1.00 | – | – | 1.00 | – | – |
| *1–49% stenosis* | 0.91 | 0.87–0.96 | <0.001 | 0.93 | 0.88–0.98 | 0.006 |
| *≥50% stenosis* | 0.83 | 0.75–0.90 | <0.001 | 0.85 | 0.78–0.94 | 0.001 |
| **Coronary artery calcium** | | n = 7,958 | | | n = 7,700 | |
| *0 Agatston units* | 1.00 | – | – | 1.00 | – | – |
| *1–99 Agatston units* | 0.93 | 0.88–0.98 | 0.008 | 0.94 | 0.89-1.00 | 0.035 |
| *≥100 Agatston units* | 0.90 | 0.83–0.96 | 0.003 | 0.93 | 0.86-1.00 | 0.050 |
| **Carotid plaque** | | n = 9,092 | | | n = 8,769 | |
| *No plaque* | 1.00 | – | – | 1.00 | – | – |
| *Unilateral plaque/s* | 0.96 | 0.91–1.01 | 0.140 | 0.98 | 0.92-1.03 | 0.382 |
| *Bilateral plaques* | 0.88 | 0.83–0.93 | <0.001 | 0.90 | 0.85–0.95 | 0.000 |

Multinomial logistic regression analysis.
a Main analysis refers to Model 2, adjusted for age at conscription, site at conscription, year at conscription, site at SCAPIS, age at SCAPIS, BMI at conscription, educational level at conscription, and physical fitness at conscription.
b The sensitivity analysis is in addition to Model 2, adjusted for smoking at conscription, height at conscription, and blood pressure at conscription.
Abbreviations: BMI, body mass index; CI, confidence interval; OR, odds ratio; SD, standard deviation; SCAPIS, Swedish CArdioPulmonary bioImage Study.

**Table S10. Odds ratios for atherosclerosis in middle age associated with each 15-unit (1 SD) increase in adolescent intelligence, with extended adjustments.**

| | Main analysis [a] | | | Sensitivity analysis with extended adjustments [b] | | | Sensitivity analysis with extended adjustments [c] | | |
|---|---|---|---|---|---|---|---|---|---|
| | Per 15-unit (1 SD) higher adolescent intelligence | | | | | | | | |
| **Outcome** | **OR** | **95% CI** | **P** | **OR** | **95% CI** | **P** | **OR** | **95% CI** | **P** |
| **Coronary stenosis** | n = 8,117 | | | n = 7,853 | | | n = 7,775 | | |
| *0% stenosis* | 1.00 | – | – | 1.00 | – | – | 1.00 | – | – |
| *1–49% stenosis* | 0.91 | 0.87–0.96 | <0.001 | 0.91 | 0.86-0.96 | <0.001 | 0.91 | 0.87-0.96 | <0.001 |
| *≥50% stenosis* | 0.83 | 0.75–0.90 | <0.001 | 0.87 | 0.80-0.94 | <0.001 | 0.85 | 0.77-0.94 | 0.001 |

Multinomial logistic regression analysis.
[a] Main analysis refers to Model 2, adjusted for age at conscription, site at conscription, year at conscription, site at SCAPIS, age at SCAPIS, BMI at conscription, educational level at conscription, and physical fitness at conscription.
[b] The sensitivity analysis is adjusted to Model 2, and participants with calcium blooming artefact on CCTA were classified as ≥50% coronary stenosis instead of 1-49% coronary artery stenosis.
[c] The sensitivity analysis is adjusted to Model 2, and participants with stent were excluded instead of being classified as ≥50% coronary stenosis.
Abbreviations: BMI, body mass index; CI, confidence interval; OR, odds ratio; SCAPIS, Swedish CArdioPulmonary bioImage Study; SD, standard deviations.

**Table S11. Odds ratios for atherosclerosis in middle age associated with each 15-unit (1 SD) increase in adolescent intelligence, according to Model 2 minus BMI at conscription.**

| | Main analysis [a] | | | Sensitivity analysis adjustments without BMI [b] | | |
|---|---|---|---|---|---|---|
| | Per 15–unit (1 SD) higher adolescent intelligence | | | | | |
| **Outcomes** | **OR** | **95% CI** | **P** | **OR** | **95% CI** | **P** |
| **Coronary stenosis** | | n = 8,117 | | | n = 8,117 | |
| *0% stenosis* | 1.00 | – | – | 1.00 | – | – |
| *1–49% stenosis* | 0.91 | 0.87–0.96 | <0.001 | 0.89 | 0.85-0.94 | <0.001 |
| *≥50% stenosis* | 0.83 | 0.75–0.90 | <0.001 | 0.80 | 0.73-0.87 | <0.001 |
| **Coronary artery calcium** | | n = 7,958 | | | n = 7,958 | |
| *0 Agatston units* | 1.00 | – | – | 1.00 | – | – |
| *1–99 Agatston units* | 0.93 | 0.88–0.98 | 0.008 | 0.91 | 0.87-0.96 | 0.001 |
| *≥100 Agatston units* | 0.90 | 0.83–0.96 | 0.003 | 0.86 | 0.80-0.92 | <0.001 |
| **Carotid plaque** | | n = 9,092 | | | n = 9,092 | |
| *No plaque* | 1.00 | – | – | 1.00 | – | – |
| *Unilateral plaque/s* | 0.96 | 0.91–1.01 | 0.140 | 0.96 | 0.91-1.01 | 0.116 |
| *Bilateral plaques* | 0.88 | 0.83–0.93 | <0.001 | 0.88 | 0.83-0.93 | <0.001 |

Multinomial logistic regression analysis.

[a] Main analysis refers to Model 2, adjusted for age at conscription, site at conscription, year at conscription, site at SCAPIS, age at SCAPIS, BMI at conscription, educational level at conscription, and physical fitness at conscription.

[b] The sensitivity analysis is according to Model 2 except BMI at conscription.

Abbreviations: BMI, body mass index; CI, confidence interval; OR, odds ratio; SD, standard deviation; SCAPIS, Swedish CArdioPulmonary bioImage Study.

**Table S12. Odds ratios for atherosclerosis in middle age associated with each 15-unit (1 SD) increase in adolescent intelligence, excluding participants with myocardial infarction, stroke, peripheral artery disease or coronary artery bypass graft.**

| | Main analysis [a] | | | Sensitivity analysis excluded participants with myocardial infarction, stroke, peripheral artery disease or coronary artery bypass graft [b] | | |
|---|---|---|---|---|---|---|
| | Per 15–unit (1 SD) higher adolescent intelligence | | | | | |
| **Outcomes** | **OR** | **95% CI** | **P** | **OR** | **95% CI** | **P** |
| **Coronary stenosis** | n = 8,117 | | | n = 7,857 | | |
| *0% stenosis* | 1.00 | – | – | 1.00 | – | – |
| *1–49% stenosis* | 0.91 | 0.87–0.96 | <0.001 | 0.92 | 0.87-0.96 | 0.001 |
| *≥50% stenosis* | 0.83 | 0.75–0.90 | <0.001 | 0.88 | 0.80-0.97 | 0.011 |
| **Coronary artery calcium** | n = 7,958 | | | n = 7,782 | | |
| *0 Agatston units* | 1.00 | – | – | 1.00 | – | – |
| *1–99 Agatston units* | 0.93 | 0.88–0.98 | 0.008 | 0.93 | 0.89-0.99 | 0.014 |
| *≥100 Agatston units* | 0.90 | 0.83–0.96 | 0.003 | 0.90 | 0.84-0.97 | 0.005 |
| **Carotid plaque** | n = 9,092 | | | n = 8,720 | | |
| *No plaque* | 1.00 | – | – | 1.00 | – | – |
| *Unilateral plaque/s* | 0.96 | 0.91–1.01 | 0.140 | 0.96 | 0.91-1.02 | 0.185 |
| *Bilateral plaques* | 0.88 | 0.83–0.93 | <0.001 | 0.89 | 0.84-0.94 | <0.001 |

Multinomial logistic regression analysis.
[a] Main analysis refers to Model 2, adjusted for age at conscription, site at conscription, year at conscription, site at SCAPIS, age at SCAPIS, BMI at conscription, educational level at conscription, and physical fitness at conscription.
[b] The sensitivity analyses are adjusted according to Model 2 and exclude participants with myocardial infarction, stroke, peripheral artery disease or coronary artery bypass graft.
Abbreviations: BMI, body mass index; CI, confidence interval; OR, odds ratio; SD, standard deviation; SCAPIS, Swedish CArdioPulmonary bioImage Study.

**Table S13. Odds ratios for atherosclerosis in middle age associated with each 15-unit (1 SD) increase in adolescent intelligence, with extended adjustments for educational level at SCAPIS.**

| | Main analysis [a] | | | Sensitivity analysis with adjustments for educational level at SCAPIS [b] | | |
|---|---|---|---|---|---|---|
| | Per 15–unit (1 SD) higher adolescent intelligence | | | | | |
| **Outcomes** | **OR** | **95% CI** | **P** | **OR** | **95% CI** | **P** |
| **Coronary stenosis** | | n = 8,117 | | | n = 7,935 | |
| *0% stenosis* | 1.00 | – | – | 1.00 | – | – |
| *1–49% stenosis* | 0.91 | 0.87–0.96 | <0.001 | 0.91 | 0.86-0.96 | 0.001 |
| *≥50% stenosis* | 0.83 | 0.75–0.90 | <0.001 | 0.85 | 0.77-0.94 | 0.001 |
| **Coronary artery calcium** | | n = 7,958 | | | n = 7,781 | |
| *0 Agatston units* | 1.00 | – | – | 1.00 | – | – |
| *1–99 Agatston units* | 0.93 | 0.88–0.98 | 0.008 | 0.92 | 0.87-0.98 | 0.007 |
| *≥100 Agatston units* | 0.90 | 0.83–0.96 | 0.003 | 0.91 | 0.84-0.98 | 0.013 |
| **Carotid plaque** | | n = 9,092 | | | n = 8,855 | |
| *No plaque* | 1.00 | – | – | 1.00 | – | – |
| *Unilateral plaque/s* | 0.96 | 0.91–1.01 | 0.140 | 0.98 | 0.92-1.04 | 0.520 |
| *Bilateral plaques* | 0.88 | 0.83–0.93 | <0.001 | 0.92 | 0.86-0.98 | 0.006 |

Multinomial logistic regression analysis.

[a] Main analysis refers to Model 2, adjusted for age at conscription, site at conscription, year at conscription, site at SCAPIS, age at SCAPIS, BMI at conscription, educational level at conscription, and physical fitness at conscription.

[b] The sensitivity analyses include additional adjustments for educational level at SCAPIS, in addition to Model 2.

Abbreviations: BMI, body mass index; CI, confidence interval; OR, odds ratio; SD, standard deviation; SCAPIS, Swedish CArdioPulmonary bioImage Study.

**Table S14. Odds ratios for atherosclerosis in middle age associated with each 15-unit (1 SD) increase in adolescent intelligence, considering data in all 11 most relevant coronary artery segments.**

| | Main analysis Considering data from any of the 11 segments [a] | | | Sensitivity analysis Considering data from all of the 11 segments [b] | | |
|---|---|---|---|---|---|---|
| | Per 15–unit (1 SD) higher adolescent intelligence | | | | | |
| **Outcome** | **OR** | **95% CI** | **P** | **OR** | **95% CI** | **P** |
| **Coronary stenosis** | n = 8,117 | | | n = 6,775 | | |
| *0% stenosis* | 1.00 | – | – | 1.00 | – | – |
| *1–49% stenosis* | 0.91 | 0.87–0.96 | <0.001 | 0.93 | 0.88-0.98 | 0.010 |
| *≥50% stenosis* | 0.83 | 0.75–0.90 | <0.001 | 0.79 | 0.71-0.88 | <0.001 |

Multinomial logistic regression analysis.

[a] Main analysis refers to Model 2, adjusted for age at conscription, site at conscription, year at conscription, site at SCAPIS, age at SCAPIS, BMI at conscription, educational level at conscription, and physical fitness at conscription.

[b] The sensitivity analysis only includes participants with data from all 11 coronary artery segments. The analysis is adjusted according to Model 2.

Abbreviations: BMI, body mass index; CI, confidence interval; OR, odds ratio; SD, standard deviation; SCAPIS, Swedish CArdioPulmonary bioImage Study.

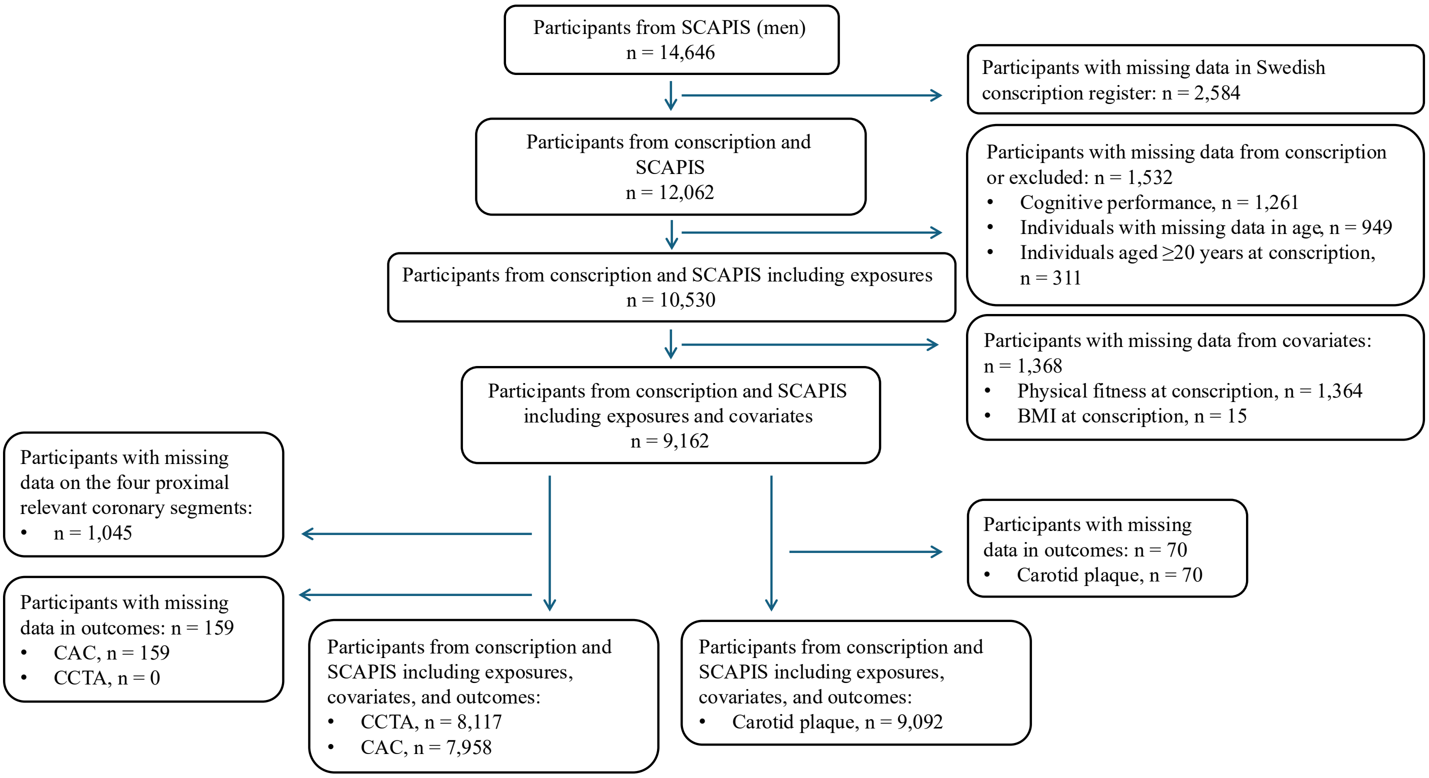

**Figure S1.** A flow chart for the study.

Abbreviations: BMI, body mass index; CAC, Coronary Artery Calcium; CCTA, coronary computed tomography angiography; SCAPIS, Swedish CArdioPulmonary bioImage Study.

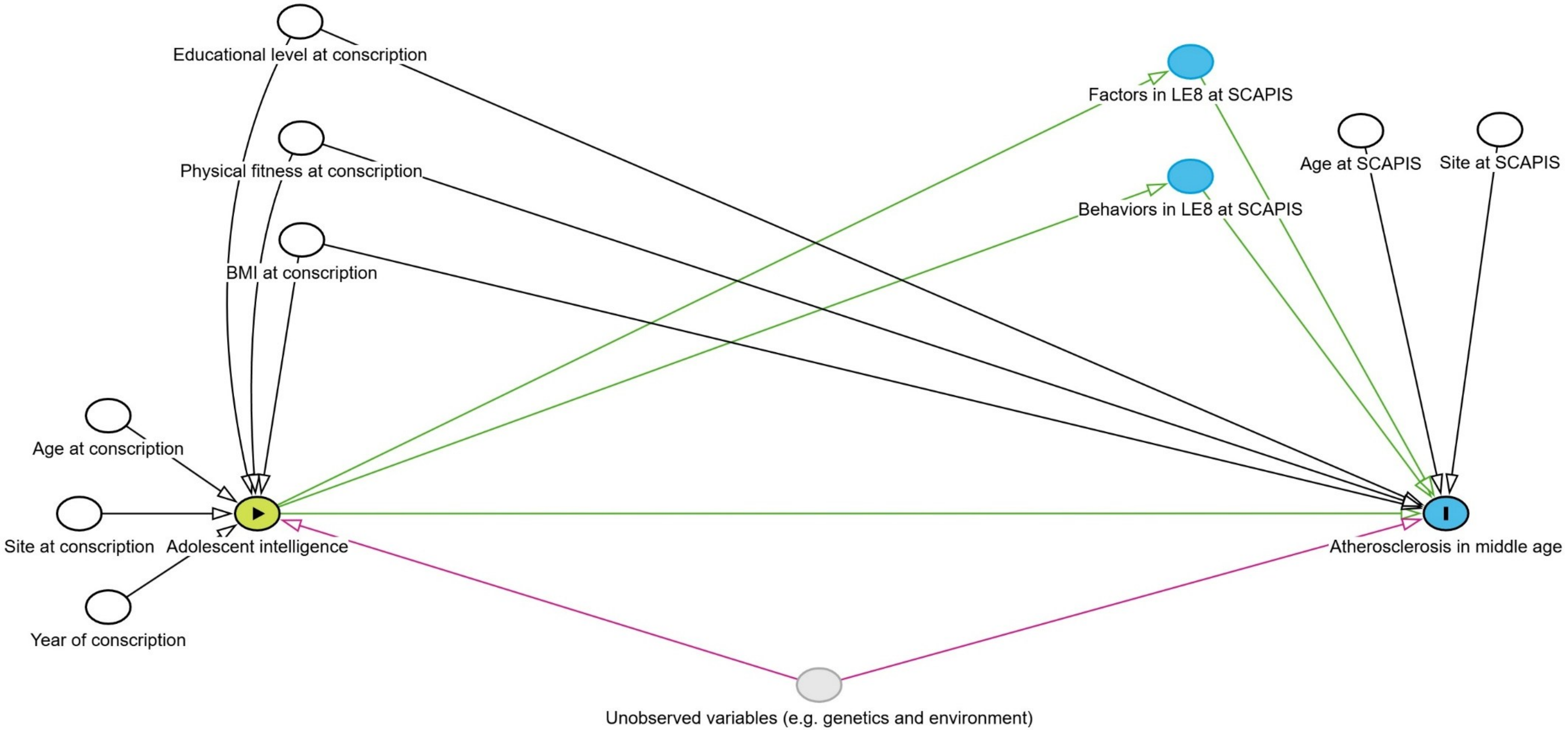

**Figure S2. Directed Acyclic Graph illustrating the association between adolescent intelligence and atherosclerosis in middle age.**

White circles represent adjusted variables, yellow circles represent the exposure, and the blue circle represents the outcome. The grey circle depicts unobserved variables. The green line indicates the causal pathway, and the pink line represents a potential biasing pathway. The factors in LE8 at SCAPIS included blood pressure, BMI, blood glucose and blood lipids. Behaviors in LE8 at SCAPIS included sleep, physical activity, smoking habits and diet.

Abbreviations: BMI, body mass index; LE8, Life´s Essential 8; SCAPIS, Swedish CArdioPulmonary bioImage Study.

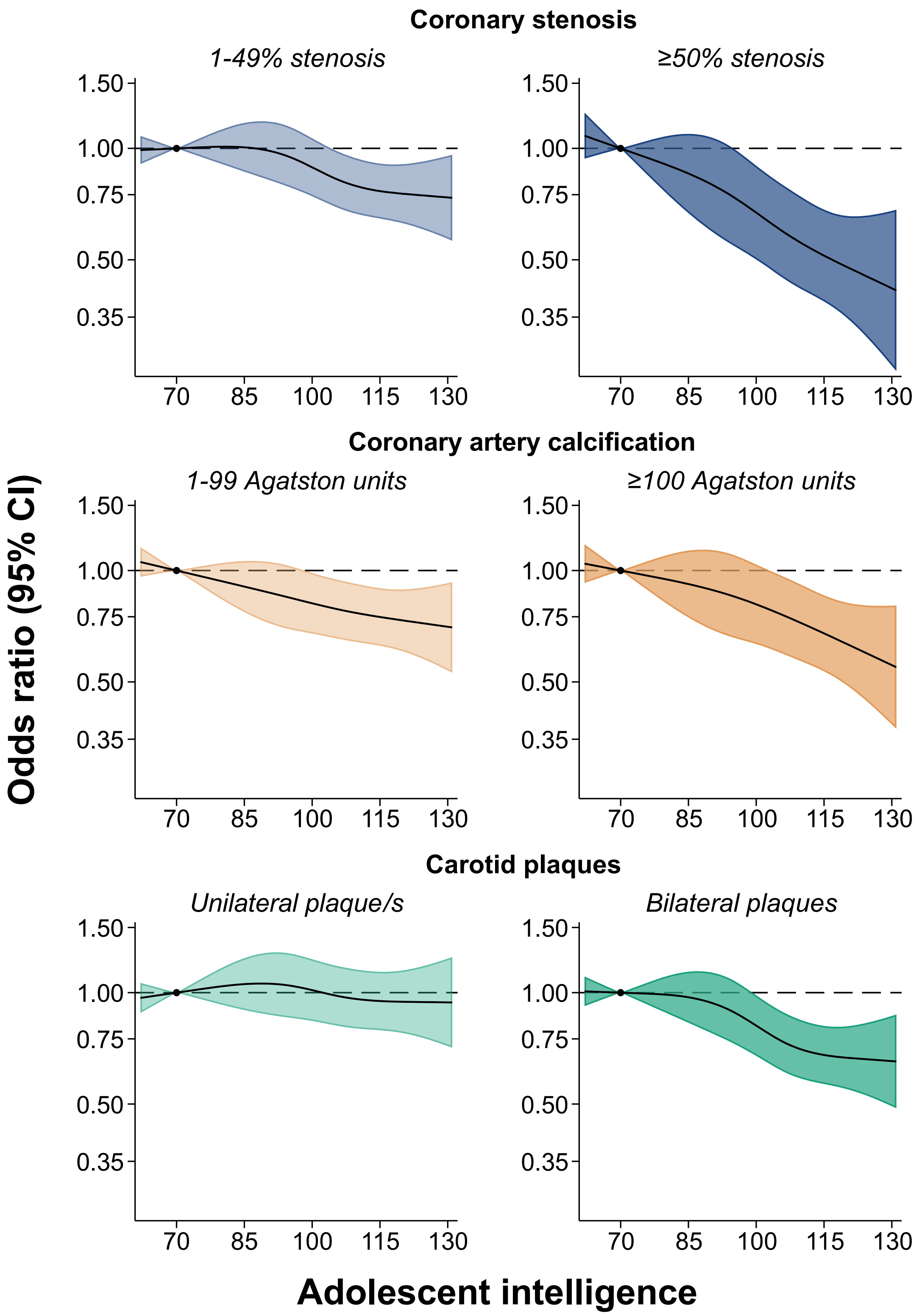

**Figure S3. Association between adolescent intelligence and coronary stenosis, CAC and carotid plaque in middle age, modeled using restricted cubic splines with multinomial logistic regression.** The reference value for adolescent intelligence is 70. The analyses are adjusted according to Model 1, age at conscription, site at conscription, year of conscription, site at SCAPIS and age at SCAPIS.

Abbreviations: BMI, body mass index; CAC, Coronary Artery Calcium; CI, confidence interval; OR, odds ratio; SCAPIS, Swedish CArdioPulmonary bioImage Study.

7. Raff GL, Chair, Abidov A, Achenbach S, Berman DS, Boxt LM, et al. SCCT guidelines for the interpretation and reporting of coronary computed tomographic

angiography. J Cardiovasc Comput Tomogr [Internet]. 2009 Mar [cited 2025 May 9];3(2):122–36. Available from: https://pubmed.ncbi.nlm.nih.gov/19272853/
